# Pleiotropy method identifies genetic overlap between orofacial clefts at multiple loci from GWAS of multi-ethnictrios

**DOI:** 10.1101/2020.11.13.20231654

**Authors:** Debashree Ray, Sowmya Venkataraghavan, Wanying Zhang, Elizabeth J. Leslie, Jacqueline B. Hetmanski, Mary L Marazita, Ingo Ruczinski, Margaret A. Taub, Terri H. Beaty

## Abstract

Based on epidemiologic and embryologic patterns, nonsyndromic orofacial clefts are commonly categorized into cleft lip with or without cleft palate (CL/P) and cleft palate alone (CP). While nearly forty risk genes have been identified for CL/P, few risk genes are known for CP. We used a new statistical method, PLACO, to identify genetic variants influencing risk of both CL/P and CP. In a combined multi-ethnic genome-wide study of 2,771 CL/P and 611 CP case-parent trios, we discovered 6 new loci of genetic overlap between CL/P and CP; 3 new loci between pairwise OFC subtypes; and 4 loci not previously implicated in OFCs. We replicated the shared genetic etiology of subtypes underlying CL/P, and further discovered loci of genetic overlap exhibiting etiologic differences. In summary, we found evidence for new genetic regions and confirmed some recognized OFC genes either exerting shared risk or with opposite effects on risk to OFC subtypes.

## INTRODUCTION

Orofacial clefts (OFCs) are the most common craniofacial birth defects that severely affect financial and psychological well-being and the overall quality of life of the affected child and their family^1^. These malformations most commonly occur as isolated defects (i.e., nonsyndromic clefts) and affect, on average, nearly 1 out of 1, 000 live births worldwide^2^. People born with OFCs require multi-displinary medical treatments; have increased risk of psychological problems^3^; have greater risk of various types of cancer (e.g., breast, brain and colon)^4^; and have increased mortality throughout the life course^5^. Overall, OFCs pose a major public health burden, with underlying biological mechanisms largely unknown.

Nonsyndromic OFCs typically manifest as a gap in the upper lip (‘cleft lip’ or CL) or the roof of the mouth (‘cleft palate’ or CP) or both (‘cleft lip and palate’ or CLP). Based on epidemiologic evidence, prevalence rates and the embryologic period when they develop, the subtypes CL and CLP are typically grouped together as the subgroup CL/P (cleft lip with or without palate)^6–8^, while CP alone forms the other subgroup. CL/P and CP have been historically analyzed separately^9–12^. While genetic studies have identified nearly 40 genetic regions (or loci) as significantly associated with risk to CL/P, fewer, around 10 loci, have been identified for CP^2,13–15^. The findings for CP have mostly been identified in the Han Chinese population^15^. Together, these genetic regions explain no more than a quarter of the estimated total heritability of risk to OFCs^16^.

Although the OFC subgroups CL/P and CP have been considered distinct, shared genetic risk variants have been suggested^9,11^. There are multiplex cleft families with both CL/P and CP present in affected relatives^17,18^. In recent years, there have been attempts to discover overlapping genetic etiology of OFC subtypes. In this context, it is important to distinguish between genetic overlap and genetic heterogeneity. While genetic heterogeneity may refer to shared genetic effects as well as subtype-specific effects (which may mean a non-null effect on one subtype and no effect on the other), genetic overlap refers to non-null genetic effects on both subtypes that may or may not be equal in magnitude and/or direction. The usual approach for identifying genetic overlap in OFCs is to compare the significant findings from one subtype with those from the other^11,13,19,20^. However, the discovery of the associated variants in the first place may be under-powered in genome-wide association studies (GWAS) of each subtype separately. For instance, success of discovery genetics has been elusive for CP, which could partially be due to smaller sample sizes of CP^19^ reflecting its lower birth prevalence. Another approach involves testing how well polygenic risk scores for one subtype can explain variation for another^13,21^, which describes overall genetic sharing, does not implicate specific regions of overlap (novel or otherwise), and may indicate lack of overlap when one subtype has a much smaller sample size^13,14^. One strategy is the ‘pooled method’ GWAS analysis^12^, where all the OFC subtypes are pooled together in a combined analysis of all OFCs. *FOXE1* has been successfully implicated as a shared risk gene using this approach^12^. While association signals from the pooled method may be driven by shared risk variants between subtypes, the pooled method does not necessarily capture only shared signals^22^, especially if sample sizes are widely different between the subtypes (e.g., the CL/P group is almost always much larger than the CP group) or if strong genetic effects exist in one group but not the other. Furthermore, if a locus is hypothesized to have opposite genetic effects on the two subtypes (e.g., *NOG*^14,19^), the pooling technique will dilute any signal and consequently will be under-powered to detect genetic overlap. Use of multi-trait methods in OFC genetic studies, such as the ones commonly used in population-based GWAS of complex traits^23–26^, is hindered by the disjoint nature of the subtypes (i.e., absence of subjects with both traits), the qualitative (binary) nature of the traits, and/or the case-parent trio design typically used to study multi-ethnic samples with OFC.

In this article, we use a new statistical method for pleiotropic analysis under composite null hypothesis, PLACO, to discover genetic variants influencing risk of the two major nonsyndromic OFC subgroups (CL/P & CP). Although PLACO was originally developed to discover pleiotropic variants between two traits from population-based studies^27^, we found it can also help identify genetic variants simultaneously influencing risk in two disease subgroups from family-based studies (see **Methods**). PLACO is particularly useful and powerful in identifying variants that increase risk of one subgroup while decreasing risk for the other, and seems to be robust to modest difference in sample sizes and in effect sizes between subgroups. We performed *a meta-analysis GWAS using PLACO on 2*, 771 CL/P and 611 CP multi-ethnic case-parent trios from the Pittsburgh Orofacial Cleft (POFC) and the Genes and Environment Association (GENEVA) studies. To dissect the genetic architecture at regions of genetic overlap between CL/P & CP, we also explored genetic overlap stratified by racial/ethnic group, investigated if the overlapping genetic etiology is modified by sex, and explored if our findings are driven by specific pairs of OFC subtypes (CL & CP or CLP & CP).

## RESULTS

### Identification of 9 loci with genetic overlap between CL/P & CP, including 2 novel loci for OFCs

At the genome-wide significance level 5 × 10^−8^, PLACO identified 1 locus in a well-recognized risk gene that also happens to be a candidate shared gene^28^, 1q32.2 (*IRF6, p* = 4.3 × 10^−12^), while 2 loci in 1p36.13 (*PAX7, p* = 6.9 × 10^−8^) and 17q22 (*NOG, p* = 6.0 × 10^−8^) are suggestive, barely missing this significance threshold (**Figure 1**). Additionally, 6 loci showed evidence for genetic overlap between CL/P & CP at a suggestive threshold of 10^−6^: 3q29 (*DLG1, p* = 5.3 × 10^−7^), 4p13 (*LIMCH1, p* = 5.0 × 10^−7^), 4q21.1 (*SHROOM3, p* = 8.1 × 10^−7^), 9q22.33 (*FOXE1, p* = 1.7 × 10^−7^), 19p13.12 (*RAB8A, p* = 6.8 × 10^−7^) and 20q12 (*MAFB, p* = 9.9 × 10^−7^). The 2 loci in *LIMCH1* and *RAB8A* are novel for OFCs. All the other genes have been implicated in GWAS of CL/P previously^2,14^, and insights into the molecular pathogenesis of OFCs via many of these genes is summarized elsewhere^29^. QQ plots from all our analyses show deviation from the null only in the tail end of the distribution of p-values (**Figures S1, S2**), indicating genetic signals rather than any systemic bias.

**Figure 1:**
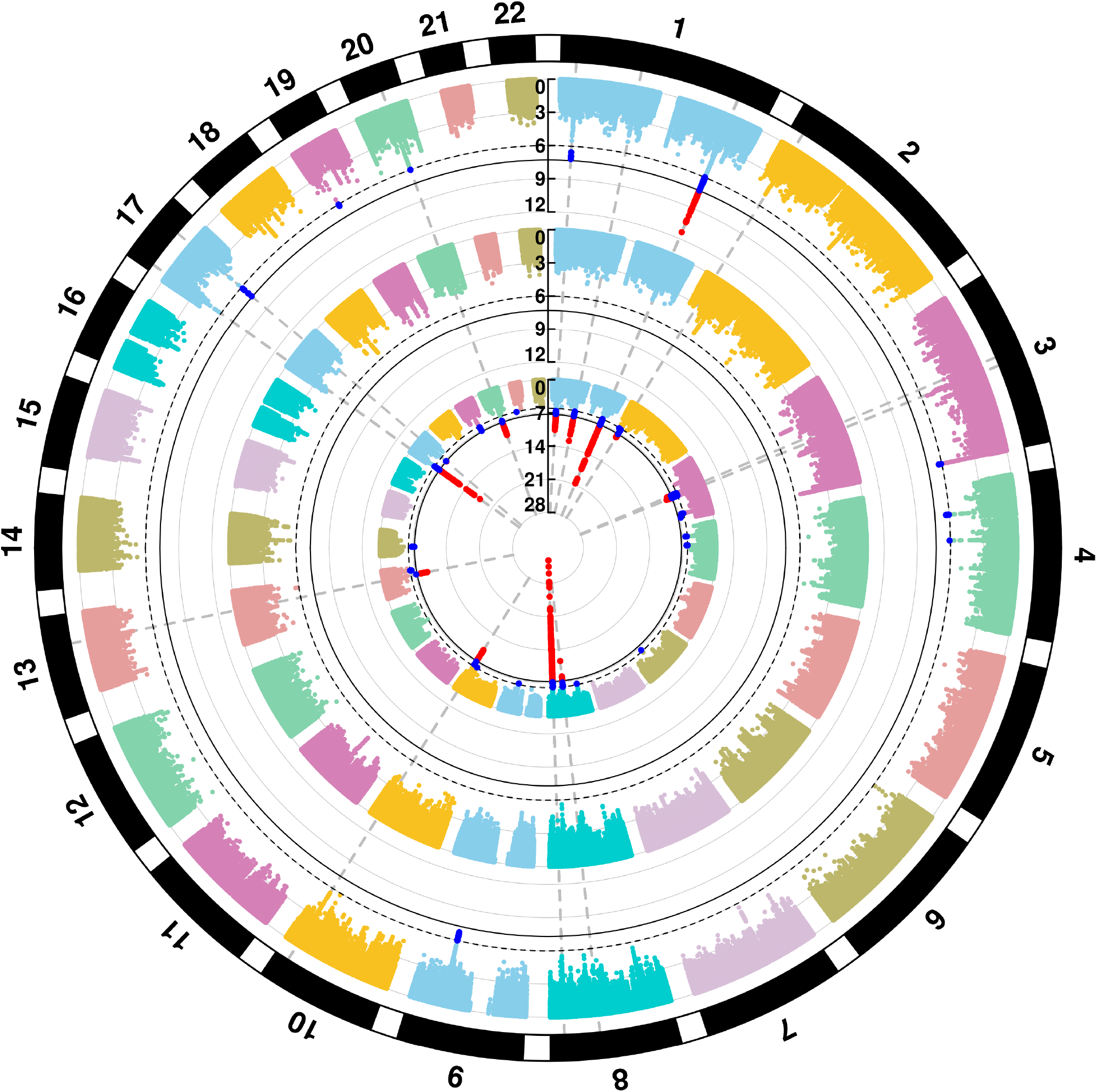
Manhattan plots for genome-wide analyses of OFC subgroups. The plots are of negative log-transformed p-values from the analysis of cleft lip with or without cleft palate (CL/P, innermost circle), of cleft palate (CP, intermediate circle), and of genetic overlap between CL/P & CP using PLACO (outermost circle). The chromosome numbers 1−22 are indicated along the outermost circumference. Solid black and dashed black circular lines are used in all plots to indicate the conventional genome-wide significance threshold 5 × 10^−8^ and a less stringent suggestive threshold 10^−6^ respectively. The variants exceeding the genome-wide and the liberal thresholds are respectively colored in red and bright blue.

### Six out of 9 loci yielding novel evidence for genetic overlap between CL/P & CP

Of the 9 loci, genetic overlap at SNPs in/near genes *IRF6*^28^, *FOXE1*^12^ *and NOG*^19^ *have been previously suggested in GWAS of clefts. We found novel, strong statistical evidence of genetic overlap at the 6 loci in/near PAX7, DLG1, LIMCH1, SHROOM3, RAB8A* and *MAFB*. In particular, PLACO provided stronger evidence for a pleiotropic association compared to the marginal association of each subtype for these markers in *DLG1, LIMCH1* and *RAB8A* loci (**Table 1**).

**Table 1:**
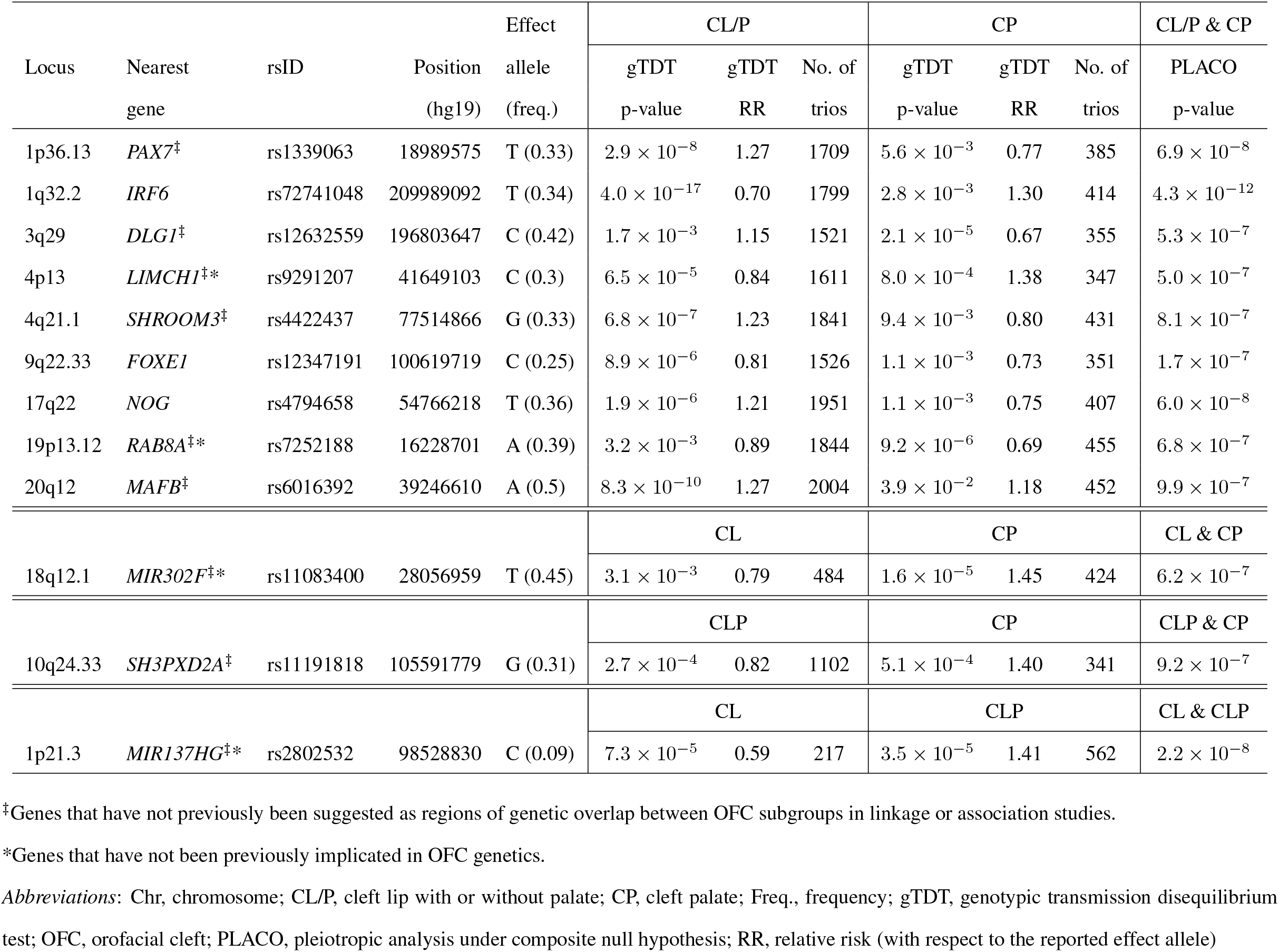
Association results for the most significant markers from the 9 loci showing statistical evidence of genetic overlap between CL/P and CP, along with 3 additional loci of genetic overlap between pairwise OFC subtypes. These loci were identified by PLACO at a suggestive threshold of 10^−6^. The genetic overlap analysis is based on all trios from both POFC and GENEVA for CL/P & CP, or CL & CP, or CLP & CP, or CL & CLP. The different types of novel genes are marked by * or ^‡^. The “No. of trios” columns give the numbers of complete informative case-parent trios as used by the gTDT method in analyzing each OFC subgroup/subtype.

### Genetic sharing at these loci are not uniform in their effect on risk

We found the chosen effect alleles at the lead SNPs in/near *FOXE1, RAB8A* and *MAFB* loci appear to increase risk for both CL/P and CP, while the effect alleles at the remaining loci affect these OFC subgroups in opposing directions as reflected by the estimated relative risks (RR) (**Table 1**). In other words, the effect alleles at the lead SNPs at *PAX7, IRF6, DLG1, LIMCH1, SHROOM3* and *NOG* seem to predispose to one OFC subgroup while protecting from the other. The estimated RRs of the top several SNPs at each of these loci further support this finding (**Figure 2**). In particular for markers in the *LIMCH1, IRF6, DLG1* and *NOG* loci, the estimated RRs of the lead SNP for subtypes CL and CLP (and hence CL/P) and their corresponding 95% confidence intervals (CIs) were all completely on the same side of the null value 1, while the estimated RR and its 95% CI for CP was completely on the other side (**Figures 3, S3, S4, S5**; panel **b**). The opposite effects of these effect alleles likely explain why these loci were not conclusively identified as influencing risk to both CL/P and CP in the ‘pooled method’ GWAS analysis of all OFC subtypes from POFC and GENEVA subjects before^12^. Additionally, the *IRF6* region appears to harbor at least 2 distinct loci: one with shared genetic effects, and another with opposite effects (**Figures 2, S6**). Evidence for both shared and opposite effects at *IRF6* has been reported previously^14,30^.

**Figure 2:**
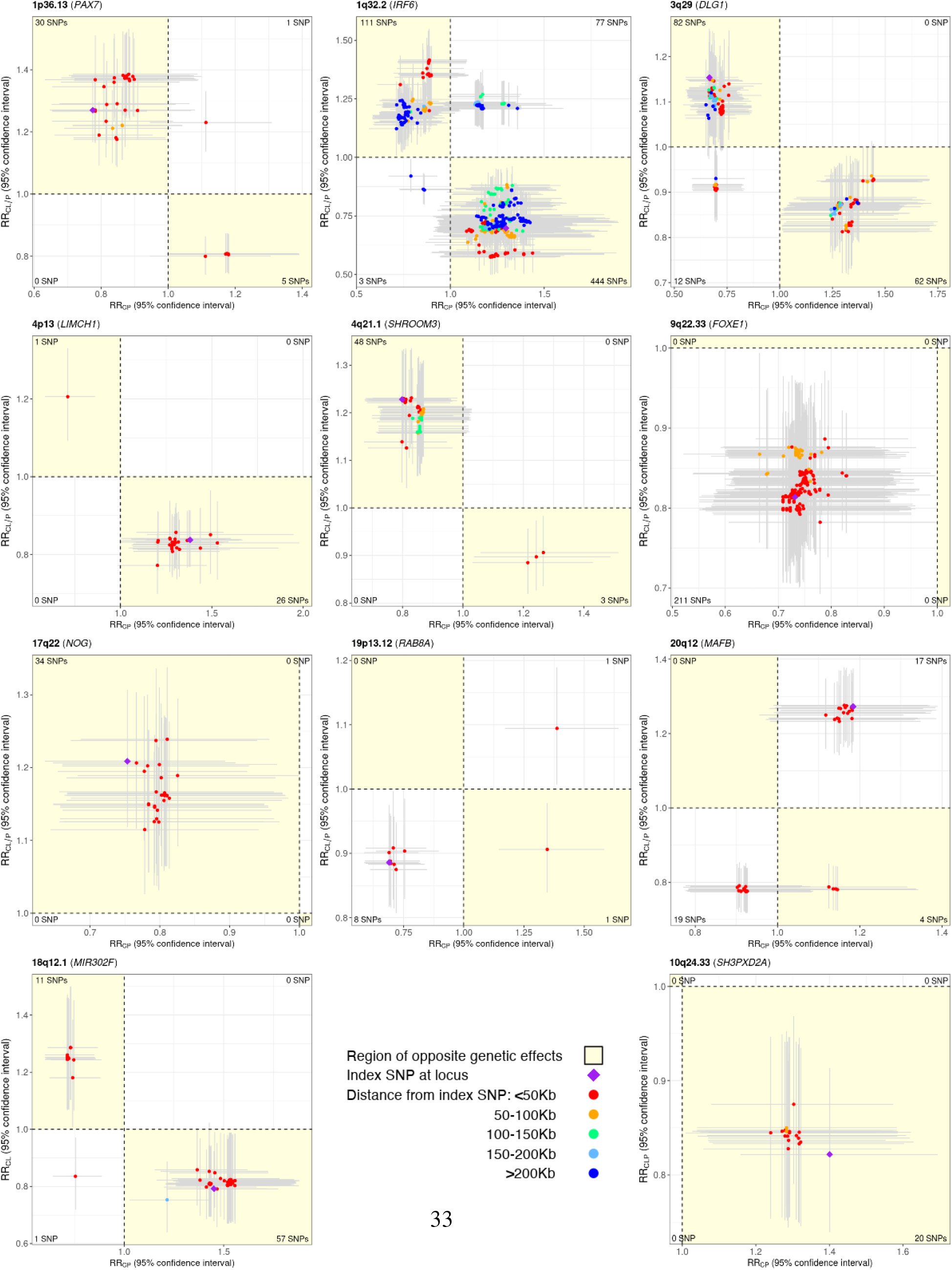
Scatter plot of relative risk (RR) estimates, along with corresponding 95% confidence intervals (CIs), for variants in the 9 loci showing statistical evidence of genetic overlap between CL/P & CP, along with 2 additional loci of genetic overlap between component OFC subtypes. RR estimates are color annotated based on distance of SNPs from the index/lead SNP. LD-based color annotation is not used since these RR estimates are from multi-ethnic analyses and consequently, there is no unique LD between SNPs. Horizontal (vertical) error bar around each RR estimate corresponds to the 95% CI for the OFC subgroup represented on the x-axis (y-axis). The region depicting opposite genetic effects of SNPs for the 2 OFC subgroups is shaded in yellow. The number of SNPs in each quadrant is printed in the corresponding corner of the plot. The SNPs plotted here are in *±*500 Kb radius and in LD *r*^2^ *>* 0.2 with the index/lead SNP, and further screened out SNPs with PLACO p-value *>* 10^−3^ from the respective genetic overlap analysis. These plots show genetically distinct etiology of CL/P and CP at 6 loci (i.e., overlapping genetic etiology with opposite effects), and of the component OFC subtypes at 2 other loci as depicted by the SNPs in the yellow shaded region.

**Figure 3:**
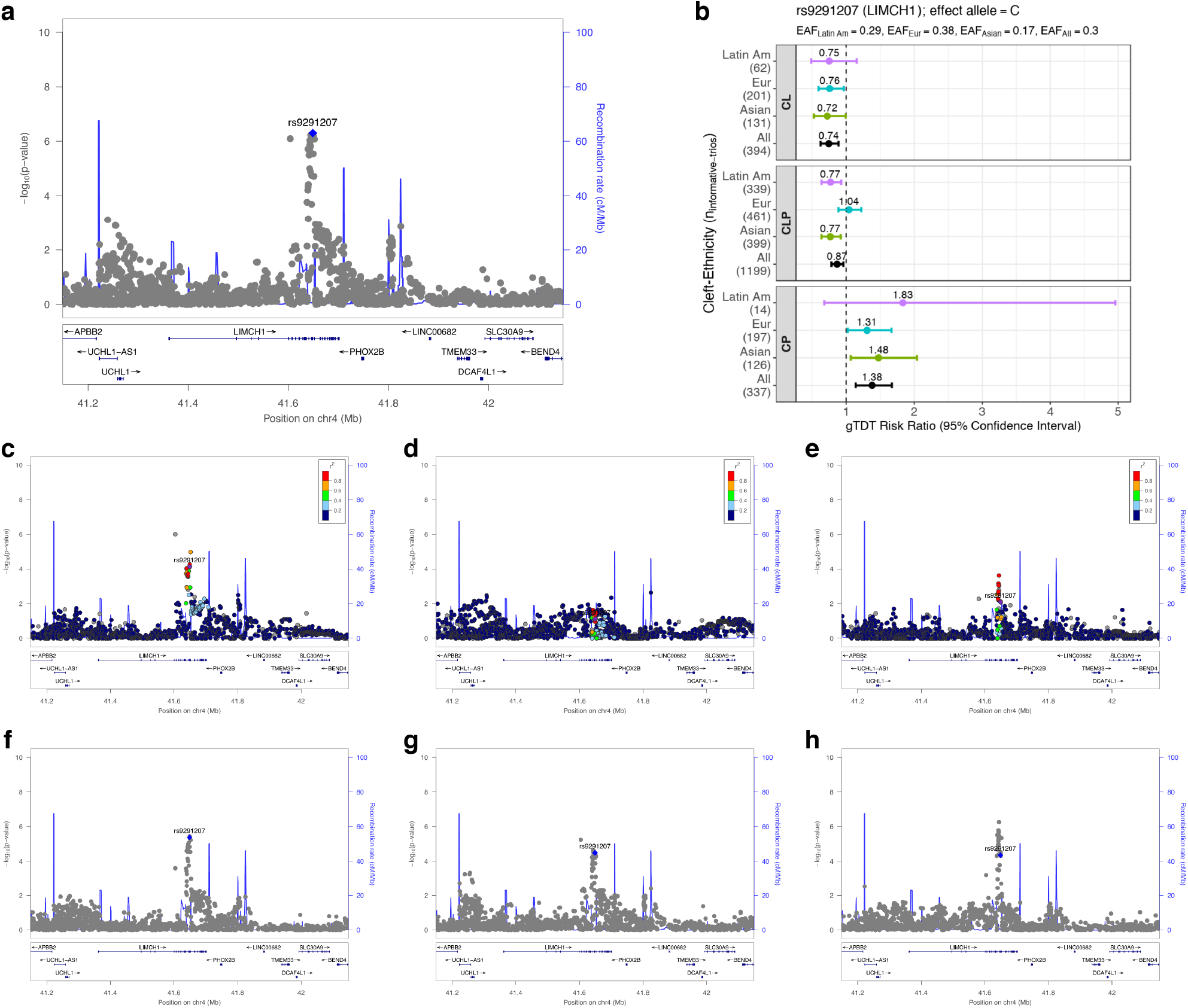
Regional association plots for 4p13 (*LIMCH1*) identified as a region of genetic overlap between CL/P & CP. LocusZoom plots focus on PLACO analysis of (**a**) CL/P & CP (multi-ethnic), (**c**) CL/P & CP in Asian ancestry, (**d**) CL/P & CP in European ancestry, (**e**) CL/P & CP in Latin American ancestry, (**f**) CL & CP (multi-ethnic), (**g**) CLP & CP (multi-ethnic), (**h**) CL & CLP (multi-ethnic). The blue or purple diamond represents the most strongly associated SNP in the region showing evidence of genetic overlap. For stratified analyses across racial/ethnic groups, the colors of the SNPs represent their LD with the lead SNP (the most strongly associated SNP from panel **a**), as shown in the color legend. For combined multi-ethnic analyses, there is no unique LD between SNPs and hence no color has been used. Panel **b** shows relative risk estimates of the lead SNP and their 95% confidence intervals as obtained from the gTDT analyses.

### Genetic overlap between subgroups CL/P & CP is consistent with overlap identified between pairwise OFC subtypes CL & CP and CLP & CP

To gain a better understanding of which subtypes are driving this evidence of genetic overlap between CL/P & CP, we applied PLACO on the pairwise component OFC subtypes (**Figure 4**). The *PAX7, SHROOM3, FOXE1* and *MAFB* loci seem to be driven by the common genetic basis of subtypes CLP & CP at these loci (**Figures S7, S8, S9, S10**; panel **g**). The rest of the loci (*LIMCH1, IRF6, DLG1, NOG* and *RAB8A*) appear to be driven by genetic overlap between CL & CP as well as CLP & CP (**Figures 3, S3, S4, S5, S11**; panels **f, g**).

**Figure 4:**
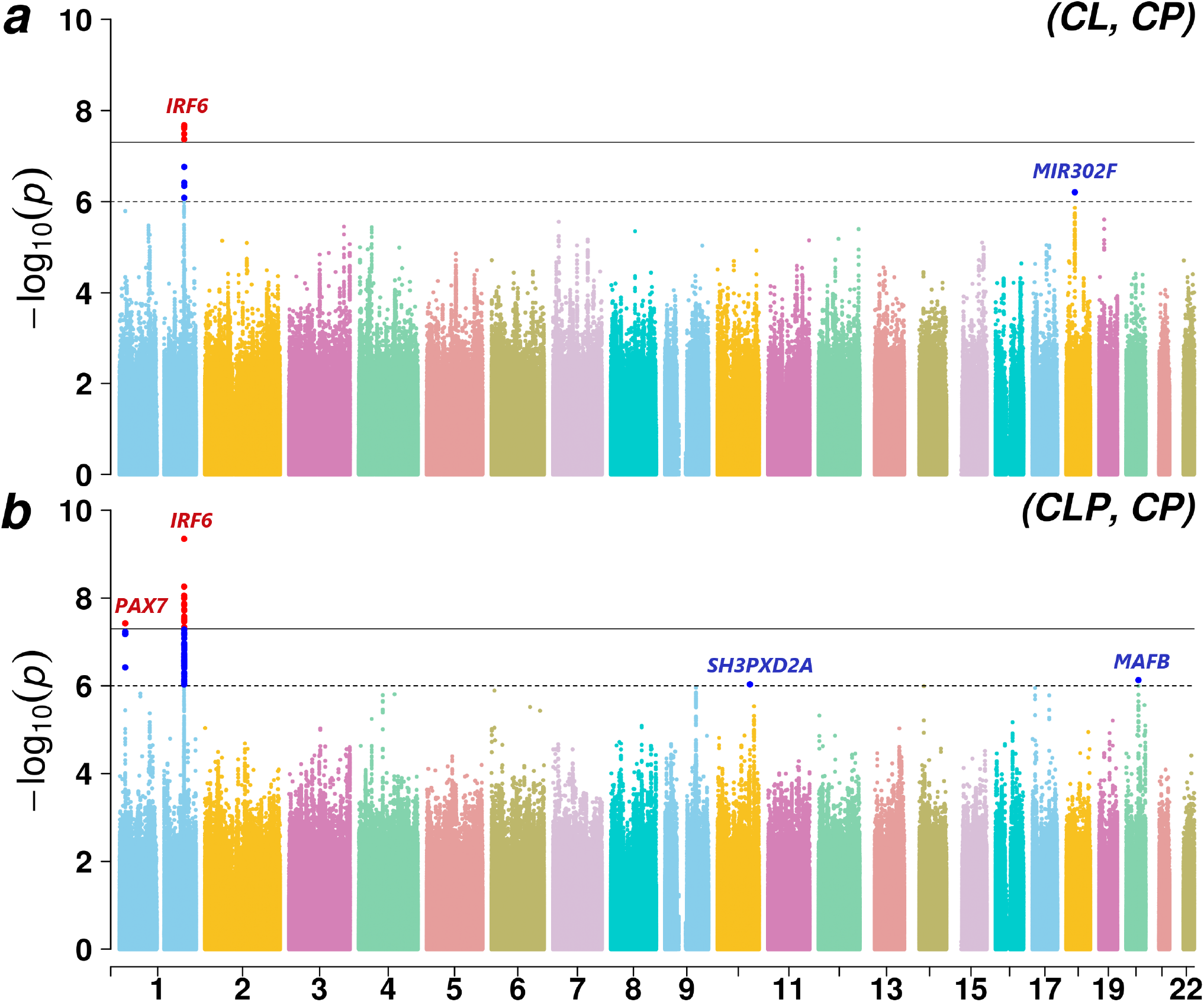
Manhattan plots for genome-wide analyses of genetic overlap between pairs of OFC subtypes: CL & CP, and CLP & CP. The chromosome numbers 1−22 are indicated along the x-axis. Solid black and dashed black lines are used in both plots to indicate the conventional genome-wide significance threshold 5 × 10^−8^ and a less stringent suggestive threshold 10^−6^ respectively. The variants exceeding the genome-wide and the liberal thresholds are respectively colored in red and bright blue.

### Identification of additional novel regions of genetic overlap between component OFC subtypes

PLACO revealed 2 loci in 18q12.1 (*MIR302F, p* = 6.2 × 10^−7^) and 10q24.33 (*SH3PXD2A, p* = 9.2 × 10^−7^) associated with CL & CP, and CLP & CP subtypes respectively at a suggestive threshold of 10^−6^ (**Table 1**). Effect alleles at the lead SNPs of both these loci increase risk for one subtype while decreasing risk for the other (**Figures S12, S13**; panel **b**). The RR estimates and their 95% CIs for the top several SNPs at these loci confirmed the opposite effect of these loci on risks of OFC subtypes (**Figure 2**). While *SH3PXD2A* has been recently implicated in the formation of CL/P in an European GWAS^31^, the *MIR302F* locus is a novel genetic risk factor for OFCs.

### Locus-specific effects at regions of genetic overlap between CL/P & CP vary by racial/ethnic group

The regional association plots seem to indicate that signals of genetic overlap at the *PAX7, FOXE1* and *NOG* loci are driven by the European subjects; *IRF6, SHROOM3* and *MAFB* loci by the Asian subjects; while the *DLG1* and *RAB8A* loci seem to draw upon evidence from both groups (**Figures S3-S11**; panels **c-e**). Similarly, evidence for the *LIMCH1* locus seems to be driven by both Asian and Latin American subjects (**Figure 3**; panels **c, e**). However, we must note that signals in these stratified analyses are confounded by differences in overall sample sizes between racial/ethnic groups (**Table S1**), sample size distribution between CL/P and CP subgroups, as well as minor allele frequency (MAF) differences across racial/ethnic groups. Consequently, the regional association plots do not fully indicate the differential information content of SNPs across these racial/ethnic groups. We, therefore, additionally provide a forest plot of RR estimates from the genotypic transmission disequilibrium test (gTDT) analyses stratified by cleft subtype and by racial/ethnic group for the lead SNP from each common locus (**Figures 3, S3-S11**; panel **b**). Notably for the Latin American subjects, the large uncertainty in the estimates for CP and the hugely skewed ratio of sample sizes between CL/P and CP are quite evident, leading to lack of power to drive signals of genetic overlap in this stratified analysis. The Asian and the European subgroups are more comparable in size, and findings from the regional association plots of these two racial/ethnic groups seem to be reflected in the forest plots as well.

### These regions of genetic overlap do not appear to be modified by sex

There is increasing evidence for sex-specific differences in human health and disorders. While CL/P is 2 times more common in males, CP is more common in females^2,29^. Recent studies have indicated sex-specific differences in pleiotropic effects on complex traits^32^. We used PLACO to test for non-null SNP×Sex interaction effects in both CL/P & CP, and failed to find any statistical evidence of pleiotropic effect modification by sex at the 9 loci of genetic overlap (**Figure S14**). We also did not find effect modification by sex at the 2 loci of genetic overlap, 18q12.1 and 10q24.33, identified between specific OFC subtypes. Tests of statistical interaction require larger sample sizes than tests of main effects; perhaps our sample size is not large enough to identify any effect modification by sex.

### Proof of principle for sensitivity of PLACO in discovering shared etiology between OFC subtypes

Analysis of subtypes CL & CLP using PLACO identified nearly all the recognized risk genes for CL/P subgroup alone as detected by the conventional gTDT analysis (**Figure 5**). In other words, the regions of genetic overlap identified by PLACO matched the shared signals captured by the pooled method analysis of CL and CLP subtypes. The RR estimates in **Table S2** indicate the effect alleles at lead SNPs in all regions of genetic overlap affect risk of both CL and CLP in the same direction, which is consistent with the vast literature of epidemiologic and genome-wide studies of OFCs^6–8,12,14,20,30^.

**Figure 5:**
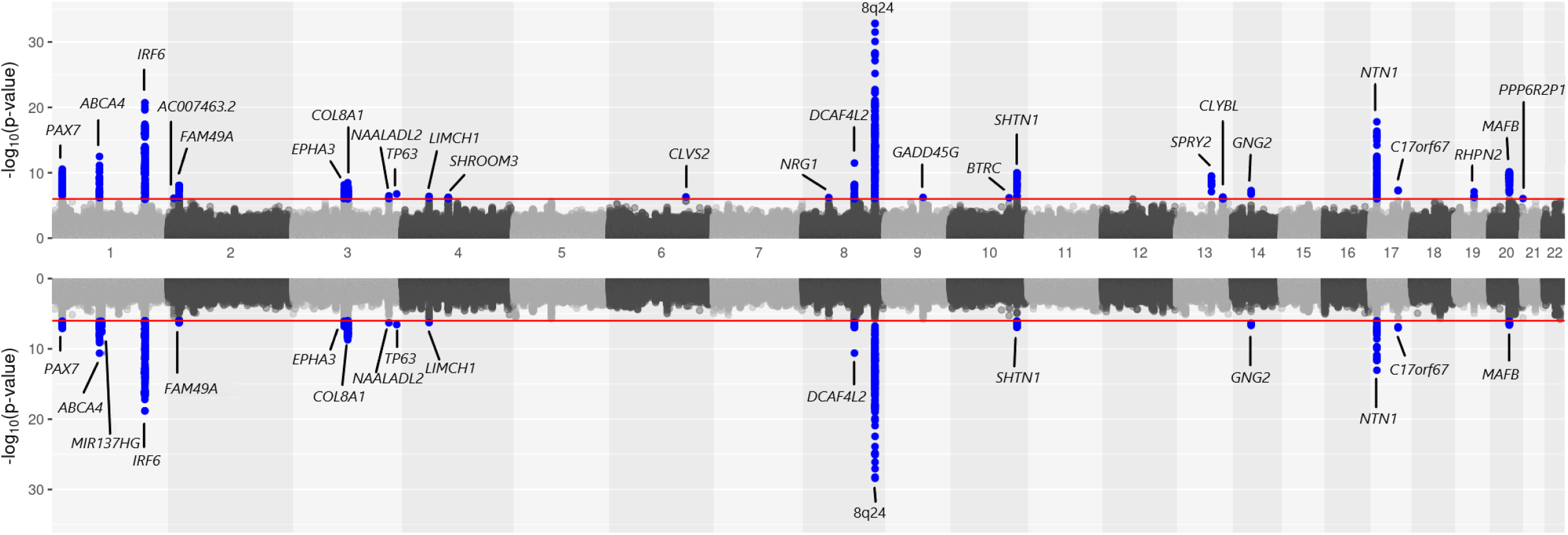
Mirrored Manhattan plot of genome-wide analyses of CL/P, and CL & CLP. Results from genetic associations of CL/P using the gTDT are shown in the upper panel. Results from shared genetic associations between CL & CLP using PLACO are shown in the lower panel. The red horizontal lines in the two panels indicate a suggestive significance threshold of 10^−6^.

### Beyond shared etiology, few loci of genetic overlap exhibit etiologic differences between CL and CLP

When investigating the RR estimates from the top several SNPs at each of the above-mentioned loci shared by CL and CLP, we found loci suggesting different pathogenesis of these subtypes at 1p22.1 (*ABCA4, ARHGAP29*) and 1q32.2 (*IRF6*) as evidenced by a large number of SNPs with opposite genetic effects (**Figure S15**). Some previous studies^14,33^ have noted differences in genetic etiologies of CL and CLP, particularly at the *IRF6* locus. Additionally, the 1p36.13 (*PAX7*), 3q12.1 (*COL8A1*), 8q21.3 (*DCAF4L2*) and 8q24 (gene desert) regions with shared effects between CL and CLP appear to have at least 2 distinct loci of genetic overlap indicated by more than one peaks (**Figure S16**). Presence of possibly independent loci in the 8q24 region has been reported previously^14^. Furthermore, PLACO revealed a novel OFC locus at 1p21.3 (*MIR137HG, p* = 2.2 × 10^−8^) with opposite genetic effects for CL & CLP at the genome-wide threshold (**Table 1** and **Figure S17 a**). The estimated RRs of the effect allele at the lead SNP of this locus indicate its protective effect on CL, and its deleterious effect on subtypes CLP and CP across racial/ethnic groups (**Figure S17 b**). This signal is, however, lost from CL/P (**Figure S17 c**) due to pooling together of opposite effects of variants on CL and CLP (**Figure S17 d, e**). Consequently, this locus near *MIR137HG* fails to show any evidence of genetic overlap between CL/P & CP (**Figure S17 f**) even though there is moderately strong statistical evidence of genetic overlap between all pairs of OFC subtypes (**Figure S17 a, g, h**).

### *In-silico* validation of robustness of PLACO to differences in sample sizes and modest subgroup-specific effects

To appreciate the advantages of PLACO and to interpret the following empirical results, it is important to briefly describe the intuition and statistical model behind PLACO. For two disease subgroups with genetic effects *β*_1_ = log(RR_1_) and *β*_2_ = log(RR_2_) for a variant, three possible situations can arise: global null, where the variant has no genetic effect on either subgroup (*β*_1_ = 0, *β*_2_ = 0); sub-null, where the variant influences risk to one subgroup but not the other (either *β*_1_ = 0, *β*_2_ ≠ 0 or *β*_1_ ≠ 0, *β*_2_ = 0); and finally non-null, where the variant influences risk to both subgroups (*β*_1_≠ 0, *β*_2_ ≠ 0). Only the non-null situation here describes genetic overlap between the two subgroups. PLACO tests a composite null hypothesis comprising both the global null and the sub-null situations, and thus rejection of this composite null provides statistical evidence of genetic overlap at a given variant (see **Methods**). On the other hand, the pooled method (previously used to identify risk variants common to both CL/P and CP)^12^ tests the global null hypothesis, and it may be rejected because either the sub-null or the non-null situations exist.

We compared PLACO with the pooled method across multiple simulation scenarios. When almost all of the simulated null variants had no effect on either OFC subgroup (Scenario I − majority of variants under the global null situation), both PLACO and the pooled method showed well-controlled type I error rates (**Figure S19 a**). As the sample sizes became skewed between the OFC subgroups, the pooled method showed inflated type I error while PLACO maintained appropriate type I error rate even at stringent error levels (**Figure S19 b-c**). When a large proportion of simulated null variants had a genetic effect on one OFC subgroup only (Scenario II − majority of variants under the sub-null situation), the pooled method had hugely inflated type I error while PLACO showed proper type I error control at stringent levels regardless of skewed sample sizes between the two OFC subgroups (**Figure S20 a-c**). This observation holds true irrespective of how widely different the MAFs are between the two simulated ethnic groups (**Figure S20 d-f**). This shows how the pooled method may show spurious signals (or increased false discoveries) if genetic effect exists in one OFC subgroup but not the other, and if there is a large sample size difference between subgroups. On the other hand, our empirical results suggest robustness of PLACO’s type I error to sample size differences between OFC subgroups; moderately strong subgroup-specific effects; and small to large MAF differences between ethnic groups.

### Empirical evidence of sensitivity of PLACO in discovering common genetic basis of OFC subgroups

We benchmarked the power of PLACO against the pooled method (even though pooled method shows increased false discoveries under the sub-null situations) along with the naive approach of declaring genetic overlap when a variant reaches genome-wide significance for the larger OFC subgroup (in our case, CL/P) and reaches a more liberal significance threshold for the other. We used two such naive approaches: one based on the criterion *p*_CL/P_ *<* 5 × 10^−8^, *p*_CP_ *<* 10^−5^ and the other *p*_CL/P_ *<* 5 × 10^−8^, *p*_CP_ *<* 10^−3^ (‘Naive-1’ and ‘Naive-2’ respectively in our figures). Regardless of the magnitude and directions of genetic effect on these OFC subgroups, and the sample size differences, PLACO showed dramatically improved statistical power to detect common genetic basis compared to the naive approaches (**Figure S21**). For instance, the ‘Naive-2’ method with a liberal threshold criterion has a 24% power, compared to 61% for PLACO, to detect simultaneous association using 1, 800 CL/P and 600 CP trios when an MAF 10% variant influences risk to both CL/P and CP with RR = 1.5. This probably explains why a genome-wide analysis of CL/P first, followed by an analysis of CP on the most significant findings, have not quite proven successful in providing evidence of overlapping association signals between these two OFC subgroups. Although PLACO was slightly less powerful than the pooled method in identifying shared risk variants, it did achieve greater power gain when detecting variants that increased risk for one OFC subgroup while decreasing risk for the other (**Figure S21**).

## DISCUSSION

In this analysis of multi-ethnic case-parent trios from the POFC and the GENEVA studies, we identified genetic overlap between nonsyndromic CL/P & CP at 1 locus in 1q32.2 reaching genome-wide significance (5 × 10^−8^), 2 loci in 1p36.13 and 17q22 barely missing this conventional threshold, and 6 loci in 3q29, 4p13, 4q21.1, 9q22.33, 19p13.12 and 20q12 yielding suggestive significance at a threshold of 10^−6^. The apparent risk SNPs at 4p13 and 19p13.12 are located in the *LIMCH1* and *RAB8A* genes respectively, which have not been implicated in OFC genetics before. We found evidence of shared etiology at 3 of these 9 loci, including the well-recognized *FOXE1* gene which influences risk to both CL/P and CP in the same direction. The effect alleles at the other loci found in this analysis appear to increase risk for one OFC subgroup but decrease risk for the other. We also identified genetic overlap between CL & CP, CLP & CP, and CL & CLP at 3 loci in 18q12.1, 10q24.3 and 1p21.3 respectively, of which the loci 18q12.1 near *MIR302F* and 1p21.3 near *MIR137HG* have not been previously shown to be associated with risk to OFCs. We replicated shared etiology of CL & CLP in/near several recognized genes, and further found opposite effects of top several SNPs at a few loci hinting at potentially different pathogenesis of these 2 OFC subtypes. None of the loci identified in this study appear to exhibit sex-dependent genetic overlap.

While *LIMCH1, RAB8A, MIR302F* and *MIR137HG* are novel risk genes for OFCs, all the other loci we identified are in/near genes previously implicated in GWAS of CL/P^2,14^. For instance, *PAX7* has been found in multiple GWAS of CL/P including a coding *de novo* variant^34^. *IRF6, FOXE1* and *NOG* have also been found in multiple GWAS, with additional evidence of functional common variants found in experimentally validated enhancer regions in these loci^12,34–36^. Association of *DLG1* with CL/P has been reported in a recent GWAS^37^ and experimentally validated in a mouse model^38^. Similarly, mouse experiments found cleft and neural tube defects (NTDs) following alterations of the *SHROOM3* gene^38^, and *MAFB* has been identified in GWAS of CLP with mouse models showing its role in palate development^29^.

The associated SNPs at the 4p13 locus occur within a topologically associating domain containing two genes: *LIMCH1* and *UCHL1*^39^. This locus was previously found to be suggestively associated with CL/P using GENEVA and POFC subjects^14^. To our knowledge, neither of these genes has been directly implicated in development of clefts to date. There is some evidence of altered methylation patterns in peripheral blood for *LIMCH1* found in Han Chinese pedigrees with children affected by NTDs^40^. Although OFCs and NTDs are considered ‘mid-line birth defects’, and supplementing mothers with folate and multivitamins during pregnancy seem to reduce risk to both^41^, it remains unknown if the same genes influence their risk. The region around the associated SNPs contains multiple putative craniofacial enhancers derived from epigenomic marks in human fetal craniofacial tissue, and the *LIMCH1* and *UCHL1* genes are decorated with marks associated with active transcription^42^. These data suggest that this locus could play a role in craniofacial development but provide no clues for the opposite effects of SNPs on the risk to CL/P and CP.

Similarly, *SH3PXD2A* was recently implicated in the etiology of CL/P for the first time in a GWAS of individuals from the Netherlands and Belgium^31^. Zebrafish and mouse models support some role of this gene in OFCs^31^. Homozygous disruption of this gene in mutant mice resulted in complete clefts of the secondary palate^43^. These studies suggest that *SH3PXD2A* might play a role in the pathogenesis of both CL/P and CP, but it is not yet clear how opposite genetic effects of the markers near this gene mechanistically influence risk to subtypes CLP and CP.

We also found opposite effects of associated SNPs for CL and CP near another novel OFC gene, *MIR302F*. There is some evidence that *MIR302* family members regulate *TP63*, a gene found mutated in ectodermal dysplasia-clefting syndrome^29^. In mouse models, members of the miR-302 family (miR-302 a-d) target different isoforms of the p63 transcription factor, the expression of which is critical for normal lip and palate development. Complete loss of p63 expression leads to CLP in mouse models^44^. Unfortunately, these studies did not include miR-302f, and little is known about the *MIR302F* gene. It is possible that *MIR302F* plays a similar critical role in craniofacial development as the other members of the *MIR302* family.

In this manuscript we have annotated the 1p21.3 and 19p13.12 loci with the genes *MIR137HG* and *RAB8A*, respectively. However, these annotations are based on proximity to the most significant SNPs and there is no specific evidence in the literature to support their role in craniofacial development. There are 3 genes in the topological domain containing the associated SNPs of 1p21.3 locus (*MIR137HG, MIR2682* and *DPYD*)^39^, which may be associated with schizophrenia and bipolar disorder^45^. *RAB8A* itself has been shown to be associated with endometrial cancer^46^. It will be an area of future work to replicate and further elucidate our findings near *RAB8A* and *MIR137HG*.

Taken together, our study provides strong statistical evidence for possible overlap in the genetic architectures of CL/P and CP. Historically, these two OFC subgroups have been thought to have distinct etiologies based on developmental origins and epidemiologic patterns. Linkage studies first identified significant evidence of linkage for markers in the *FOXE1* region in CL/P multiplex families^47^; subsequent studies confirmed this gene as a risk factor for both CL/P and CP^11,48^. Linkage analysis identified a susceptibility locus near *TBX22* for CL/P and a later study found mutations in *TBX22* in CP individuals^17^. Fine-mapping of translocation breakpoints revealed an important role of *SATB2* in cases with CP; a few years later, a candidate gene study identified significant association of a variant in *SATB2* with CL/P in two Asian populations^10,17^. There exists some evidence that variants in *IRF6*^28^, *GRHL3*^28^, *ARHGAP29*^49–51^ *and MSX1*^17^ regions may affect risk to both CL/P and CP in the same direction (often termed as ‘shared genetic risk’). Note, much of this evidence are from patterns of Mendelian inheritance of rare variants in extended pedigrees; genetic overlap of common variants in nonsyndromic OFCs may not follow the same patterns. In recent GWAS, variants near *IRF6*^20^ and *NOG*^14,19^ have shown weak evidence of decreased risk for one OFC subgroup and increased risk for the other. Among these well-recognized risk genes for OFCs, only *FOXE1* has been successfully implicated as a shared risk gene in GWAS^12^. Our method PLACO not only replicated this finding for *FOXE1*, but also provided strong statistical evidence for genetic overlap at *IRF6* and *NOG*. PLACO found no evidence of genetic overlap between CL/P & CP at common variants (MAF ≥5%) in/near *SATB2* (chr2:200134004-200316268, including rs6705250 and rs12105015; *p*_min_ = 0.03), *GRHL3, ARHGAP29* or *MSX1* (**Figure S18**). It is possible that rarer variants drive genetic overlap in these regions, and PLACO is currently only applicable to common variants.

In summary, this study advanced our knowledge of the genetic architecture controlling risk to OFCs by enriching the current inventory of OFC-associated genes with novel genes possibly driving common genetic basis of OFC subtypes. Lack of granularity of cleft subtype in animal models precludes experimental validation of our findings. No bioinformatics analysis on existing large-scale databases is equipped to explain how some of these loci could affect the two OFC subgroups in opposing directions. Instead we utilized *in-silico* validation techniques such as statistical simulation experiments, sensitivity analyses, and proof-of-principle analysis. Our extensive *in-silico* validation showed PLACO’s robustness to subgroup-specific effects (not a situation of genetic overlap), population-specific differences in MAF, and sample size differences between OFC subgroups. Our proof-of-principle analysis of subtypes CL & CLP using PLACO and replicating those findings with genetic associations of subgroup CL/P emphasized the shared etiology successfully identified by PLACO. More granular functional studies than those currently available are needed to clearly understand the differences in how some of the genes identified here could affect risk of one subtype versus another.

## METHODS

### The POFC and the GENEVA studies

Case-parent trios ascertained through cases with an isolated, nonsyndromic OFC in the GENEVA study were largely recruited through surgical treatment centers by multiple investigators from Europe (Norway and Denmark), the United States (Iowa, Maryland, Pennsylvania, and Utah) and Asia (People’s Republic of China, Taiwan, South Korea, Singapore, and the Philippines)^12,49^. Type of cleft, sex, race as well as common environmental risk factors were obtained through direct maternal interview^49^. The research protocol was approved by the Institutional Review Boards (IRB) at the Johns Hopkins Bloomberg School of Public Health and at each participating recruitment site. Written informed consent was obtained from both parents and assent from the child was solicited whenever the child was old enough to understand the purpose of the study.

The POFC study included case-parent trios ascertained through a proband with an isolated nonsyndromic CL/P or CP from multiple populations, and a large number of OFC cases and ethnically matched controls from some of these same populations^12,52^. We, however, used only the case-parent trios from POFC in this study. Similar information about type of cleft, sex, race and common environmental risk factors were collected through direct maternal interview. The research protocol was approved by the IRB at the University of Pittsburgh and all participating institutions, and informed consent was obtained from all participants.

The distribution of trios by cleft subtype and racial/ethnic groups from both studies is given in **Table S1**. It is to be noted that originally 412 individuals from POFC were included in GENEVA^52^; we have subsequently removed them from our GENEVA dataset to avoid duplication. Thus, in this article, these two studies represent independent, non-overlapping case-parent trios from three major racial/ethnic groups (European, Asian, and Latin American). Instead of having separate discovery and replication samples, we decided to combine the two studies, which should have improved power to detect genetic associations over a two-stage discovery-replication approach^53^.

### Genotyping, imputation and quality control

Participants in the GENEVA study were genotyped on the Illumina Human610 Quadv1 B array with 589,945 SNPs at the Center for Inherited Disease Research (https://cidr.jhmi.edu/). We re-imputed this dataset using the Michigan Imputation Server^54^ to take advantage of more efficient imputation tools and more recent, larger reference panels. Before imputing, we dropped SNPs with MAF*<*1%, performed trio-aware phasing of the haplotypes from the observed genotypes using SHAPEIT2^55^, and original genotyped SNPs on build hg18 were lifted over to hg19 (http://github.com/sritchie73/liftOverPlink). We used the 1000 Genomes Phase 3 release 5 reference panel for imputation. Note, trio-aware phasing before imputation is critical; ignoring the family information may lead to biased downstream results^56^. ‘Hard’ genotype calls were made by setting threshold 0.1 within PLINK 2.0 (www.cog-genomics.org/plink/2.0/)^57^. If the calls have uncertainty *>*0.1 (i.e., genotype likelihoods *<*0.9), they were treated as missing; the rest were regarded as hard genotype calls. We took the following quality control measures: all genotyped SNPs with missingness *>*5%, Mendelian error rate *>*5%, all SNPs with MAF*<*5% as well as those showing deviation from Hardy-Weinberg equilibrium (HWE) at *p <* 10^−4^ among parents were dropped. All imputed SNPs were filtered to exclude any with *R*^2^ *<* 0.3 using BCFtools-v1.9 (https://samtools.github.io/bcftools). Additionally, individuals with low genotype information or evidence of low-quality DNA, individuals with SNP missingness *>*10%, and individuals duplicated across the POFC and GENEVA datasets were excluded. Only complete trios were kept for the final analysis. The final GENEVA dataset contained 6, 762, 077 autosomal SNPs, including both observed and imputed SNPs having MAF ≥5% among parents, for 1, 939 complete case-parent trios.

The case-parent trios from the POFC study were genotyped on 539,473 SNPs on the Illumina HumanCore + Exome array. For imputation, data were phased with SHAPEIT2 and imputed using IMPUTE2^58^ to the 1000 Genomes Phase 3 reference panel as described previously^52^. Incomplete trios, trios with parents from different racial/ethnic groups and racial/ethnic groups with insufficient sample sizes for effective imputation were dropped. The same quality control measures as GENEVA were used to remove rare and poor quality SNPs. The final POFC dataset contained 6, 350, 243 autosomal SNPs, including both observed and imputed SNPs having MAF ≥5% among parents, for 1, 443 complete case-parent trios.

We meta-analyzed the POFC and the GENEVA studies (which are independent) to increase sample size and power. All SNPs with mismatch in allele or base pair information (hg19) between the two studies were removed. The final meta-analyzed dataset contained 6, 761, 961 SNPs, which includes all SNPs common to the two studies and SNPs unique to any one study.

### PLACO: pleiotropic analysis under a composite null hypothesis

Consider genome-wide studies of two disorders *Y*_1_ and *Y*_2_ based on *n*_1_ and *n*_2_ case-parent trios respectively who were genotyped/ imputed or sequenced at *p* SNPs. For a given SNP, assume an additive genetic model where the relative risks associated with the two disorders are RR_1_ and RR_2_. The corresponding genetic effect parameters are respectively *β*_1_ = log(RR_1_) and *β*_2_ = log(RR_2_). One may assume any other genetic inheritance model, and the following is still applicable. In each study, the genotypic transmission disequilibrium test (gTDT) using a conditional logistic framework^59,60^ may be used to obtain the maximum likelihood estimates 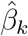 and its standard error 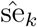 which are used to construct the summary statistic 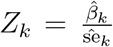 for *k* = 1, 2. Since TDTs for trios protect against confounding due to population stratification, one may combine multi-ethnic case-parent trios when analyzing the two disorders.

To implement PLACO, we start with the summary statistics *Z*_1_ and *Z*_2_ from the two disorders across all SNPs genome-wide. For practical purposes, we assume the datasets for the two disorders are independent since case-parent trios are ascertained based on the disease status of the child and it is unlikely to have subjects shared between the two datasets. While the usual multi-trait methods^23,24^ test the null hypothesis of no association of a given SNP with any disorder (i.e., *β*_1_ = 0 = *β*_2_) against the alternative hypothesis that at least one disorder is associated, PLACO tests the composite null hypothesis that at most one disorder is associated with the SNP against the alternative that both disorders are associated^27^. Mathematically, PLACO tests

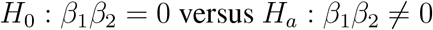

so that rejection of the null hypothesis statistically indicates genetic overlap between disorders. The null hypothesis *H*_0_ is a composite of the global null *{β*_1_ = 0 = *β*_2_*}*, and the sub-nulls *{β*_1_ = 0, *β*_2_ ≠ 0*}* and *{β*_1_ ≠ 0, *β*_2_ = 0*}*. Suppose, across the genome, the global null holds with probability *π*_00_ under which the summary statistics *Z*_1_ and *Z*_2_ have asymptotic standard normal distributions. Further assume that the first sub-null holds with probability *π*_01_ where *Z*_1_ has a standard normal distribution and *Z*_2_ has a shifted normal distribution *N* (*µ*_2_, 1). For a given disorder, the relative risks of SNPs with a non-null effect vary genome-wide. Consequently, there is no fixed value that the mean parameter *µ*_2_ takes, and to capture this variability in effect sizes we assume a random effect on the mean − a 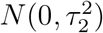 distribution. Similarly, assume that the second sub-null holds with probability *π*_02_ and *Z*_2_ *∼ N* (0, 1) while *Z*_1_ given *µ*_1_ has a *N* (*µ*_1_, 1) distribution, where *µ*_1_ is assumed to follow a 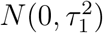 distribution.

Thus, under the composite null hypothesis *H*_0_, PLACO assumes (a) *Z*_1_ and *Z*_2_ are independent *N* (0, 1) variables when *{β*_1_ = 0, *β*_2_ ≠ 0*}* holds; (b) *Z*_1_ and *Z*_2_ are independent *N* (0, 1) and 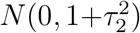 variables respectively when *{β*_1_ = 0, *β*_2_ ≠ 0*}* holds; and (c) *Z*_1_ and *Z*_2_ are independent 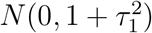 and *N* (0, 1) variables respectively when *{β*_1_ ≠ 0, *β*_2_ = 0*}* holds. We have described the rationale and other considerations behind this choice of PLACO model previously^27^. The PLACO test statistic is

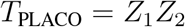

and its approximate, asymptotic p-value is given by

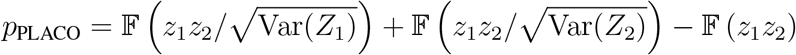

where *z*_1_ and *z*_2_ are the observed *Z*-scores for the two disorders at any given SNP; 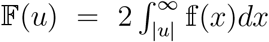 is the two-sided tail probability of a normal product distribution at value *u*; and Var(*Z*_1_) and Var(*Z*_2_) are the estimated marginal variances of these *Z*-scores under the above distributional model^27^. Open-source implementation of PLACO in R^61^ is available at https://github.com/RayDebashree/PLACO. While PLACO was originally proposed for two traits from population-based studies (e.g. case-control traits)^27^, here we showed PLACO can very well be used for family studies (**Supplementary S2**) as long as the summary statistics are obtained after appropriately accounting for all confounding effects, including relatedness and population stratification.

### Statistical analyses

For all analyses presented here, we focused on bi-allelic SNPs with MAF≥5%, where MAF is calculated based on only the parents (founders) using PLINK 2.0. For each study separately, we obtained summary statistics of genetic association between each variant and each OFC subgroup using the gTDT model for case-parent trios under an additive genetic model as implemented in R package `trio`^59^ (v3.20.0). We used the gTDT over the allelic TDT because of its several advantages^62^: gTDT can be more powerful; yields parameter estimates, standard errors along with p-values; and enables direct assessment of RR. However, unlike the allelic TDT^63^, the gTDT assumes a specific mode of inheritance; here we chose an additive model. The gTDT in `trio` package uses the minor allele of the input dataset as the effect allele. Since the gTDT is applied on each subgroup and each study separately, it is possible that a minor allele is not the same across subgroups and across studies. Therefore, we set the minor allele from the CL/P subgroup in POFC as the effect allele for all analyses. We, then, meta-analyzed the gTDT results over the two studies using inverse-variance weighted fixed effects meta-analysis. We implemented PLACO on the meta-analyzed gTDT results from subgroups CL/P and CP to identify possible genetic overlap between them. To identify regions of significant genetic overlap, we used the conventional genome-wide threshold 5 × 10^−8^ and also a suggestive threshold of 10^−6^.

We explored if any identified region of genetic overlap was modified by sex. To do this, we first obtained summary statistics from the 1 df SNP×Sex analysis using the gene-environment gTDT model in trio package (again assuming additive genetic model); then meta-analyzed the SNP×Sex estimates across the two studies; and finally applied PLACO on the meta-analyzed 1 df SNP×Sex summary statistics. For each analysis, we created Manhattan plots to show signals, and QQ plots to check for potential bias in the association results. For the QQ plots, we also calculated the genomic inflation factors at the 50^th^ percentile (*λ*_0.5_) to quantify the extent of the bulk inflation, and at the 1/10^th^ of a percentile (*λ*_0.001_) to quantify inflation towards the meaningful tail of the distribution. We took the p-values from a given method (e.g., gTDT, PLACO), mapped them to 1 df *χ*^2^ statistics, and calculated *λ*_*x*_ as the ratio of empirical 100(1 *−x*)^th^ percentile of these statistics and the theoretical 100(1 *− x*)^th^ percentile of 1 df *χ*^2^ distribution.

### Stratified analyses

We considered two stratified analyses: one stratified by racial/ethnic group and the other by cleft subtype (CL, CLP, CP). As described before, genome-wide meta-analyzed gTDT summary statistics were obtained for CL/P and for CP within each of the three major racial/ethnic groups: European, Asian and Latin American. PLACO was applied on CL/P and CP summary data within each racial/ethnic group. For the OFC subtype stratified analyses, meta-analyzed gTDT summary statistics were obtained for each cleft subtype, and then PLACO was applied on each of the three pairwise combinations to compare and contrast results against those from the main analysis.

### Locus annotation and candidate gene prioritization

For each analysis of genetic overlap, we defined independent loci by clumping all the suggestively significant SNPs (*p*_PLACO_ *<* 10^−6^) in a *±*500 Kb radius and with linkage disequilibrium (LD) *r*^2^ *>* 0.2 into a single genetic locus. This clumping was done using `FUMA`^64^ (`SNP2GENE` function, v1.3.5e). Since we performed multi-ethnic analysis, we separately used 1000G Phase 3 EUR, EAS and AMR as reference populations for LD calculation. The number of independent loci and the index SNP for each locus (chosen to be the most significant SNP) were the same regardless of the racial/ethnic groups assumed for LD calculation. To define the bounds of each locus, as used in the regional association plots annotated by effect size directions (**Figures S6, S16** and **S17 e**), we took the minimum of lower bounds and the maximum of upper bounds across racial/ethnic groups. We mapped each locus to the gene nearest to the lead SNP using `FUMA`. We used LocusZoom^65^ to get regional association plots with gene tracks that allowed us to examine detailed evidence of association at each identified locus. For these LocusZoom plots, we used genome build hg19 with no specified LD reference panel due to the multi-ethnic nature of our analysis. The LocusZoom plots for the stratified analyses of the three racial/ethnic groups, however, use the corresponding LD reference panel.

### Validation

As mentioned before, we do not have separate discovery and validation samples; combining samples improves power to detect genetic associations over a two-stage discovery-replication approach^53^. Experimental validation in animal models is not possible due to the lack of granularity of cleft subtypes in mouse or zebrafish models. No current bioinformatics analysis can fully explain the opposite effects of the loci discovered here (where the effect allele increases risk for one OFC subgroup but decreases risk for another). To provide confidence on PLACO’s findings, we undertook three complementary approaches: (1) a proof-of-principle analysis of subtypes CL & CLP using PLACO and matching those findings with genetic associations of subgroup CL/P to emphasize the shared etiology that PLACO successfully identified; (2) an *in-silico* validation of PLACO using simulated data; and (3) an assessment of our findings based on existing literature. In particular, our extensive empirical validation involves showing (i) PLACO’s robustness to subgroup-specific effects, population-specific differences in MAF, and sample size differences between OFC subgroups; and (ii) massive power gains achieved by PLACO in detecting genetic overlap (whether shared or in opposing directions) compared to other commonly-used variant-level approaches.

### *In-silico* evaluation of PLACO using simulated data for OFC trios

We simulated two bi-ethnic case-parent trio datasets with a total of 2, 400 trios mimicking independent studies of CL/P and CP. We assumed, without loss of generality, that the two ethnic groups have equal sample sizes for a particular OFC subgroup, and considered situations where the OFC subgroups either have comparable (1:1) or unbalanced (3:1) or largely unbalanced (7:1) sample sizes similar to what we saw for the POFC and the GENEVA studies. We simulated the two ethnic groups such that they are different in terms of OFC subgroup prevalence, and in terms of MAF at any given variant. We compared type I error and power of PLACO with the pooled method and the naive approach of declaring genetic overlap when a variant reaches genome-wide significance for the subgroup with the larger sample size and reaches a more liberal significance threshold for the second subgroup. See **Supplementary S2** for more details on our simulation experiments.

### Data availability

The POFC and the GENEVA studies are publicly available on dbGaP (https://www.ncbi.nlm.nih.gov/gap/, study accession numbers phs000774.v1.p1 and phs000094.v1.p1).

### Genome build

All genomic coordinates are given in NCBI Build 37/UCSC hg19.

## Data Availability

This work is entirely on existing genetic data from the POFC and the GENEVA studies, which are publicly available on dbGaP
(https://www.ncbi.nlm.nih.gov/gap/, study accession numbers phs000774.v1.p1 and phs000094.v1.p1).

## Web Resources

The R source code for the genetic overlap test `PLACO` can be found in GitHub (https://github.com/RayDebashree/PLACO/). R `trio` package can be found in Bioconductor (https://bioconductor.org/packages/release/bioc/html/trio.html).

## Supplemental Data

Supplementary information is available on the journal website.

## Acknowledgments

This research was supported in part by the NIH grants R03DE029254 (D.R., S.V., J.B.H., T.H.B.), R03DE027121 (S.V., W.Z., M.A.T., T.H.B.), R00DE025060 (E.J.L.), R01DE016148 (M.L.M.) and U24OD023382 (D.R.). It was carried out using computing cluster - the Joint High Performance Computing Exchange - at the Department of Biostatistics, Johns Hopkins Bloomberg School of Public Health.

## Author Contributions

D.R., M.A.T. and T.H.B. designed this study and performed overall project management. W.Z. and J.B.H. performed imputation and processing of the GENEVA data. D.R., S.V. and W.Z. analyzed the POFC and the GENEVA data. D.R., E.J.L., I.R., M.A.T. and T.H.B. interpreted the scientific findings. D.R. and S.V. performed statistical simulation experiments. M.L.M. and T.H.B. provided the POFC and the GENEVA data respectively. D.R. drafted the initial manuscript. All authors reviewed and approved the final manuscript.

## Competing Financial Interests

The authors declare no competing financial interests.

## Correspondence

Correspondence should be addressed to D.R. or T.H.B.

## Notes

### Competing Interest Statement

The authors have declared no competing interest.

### Author Declarations

This manuscript involves data analysis of only existing de-identified genetic studies that are publicly available on dbGaP. There is no new data collection or patient contact. So, there is no IRB associated with this work. The research protocols of the original studies were approved by IRB at Johns Hopkins Bloomberg School of Public Health, at the University of Pittsburgh, and at each participating recruitment site. The details are provided in the Methods section of this manuscript.

