## Supplemental Data for "Pleiotropy method identifies genetic overlap between orofacial clefts at multiple loci from GWAS of multi-ethnictrios"

### Supplementary S1

#### Additional tables and figures for the genetic overlap analysis of clefts.

**Table S1:** Distribution of independent complete case-parent trios in the POFC and the GENEVA studies by racial/ethnic group and by cleft subtypes.

| Study | Ethnicity | Sex* | CL | CLP | CP | All OFCs |
|---|---|---|---|---|---|---|
| POFC | All | All | 263 | 1021 | 159 | 1443 |
| | | Female | 104 | 365 | 95 | 564 |
| | | Male | 159 | 656 | 64 | 879 |
| | Asian | All | 85 | 199 | 38 | 322 |
| | European | All | 89 | 314 | 93 | 496 |
| | Latin American | All | 89 | 508 | 28 | 625 |
| GENEVA | All | All | 431 | 1056 | 452 | 1939 |
| | | Female | 186 | 348 | 254 | 788 |
| | | Male | 245 | 708 | 198 | 1151 |
| | Asian† | All | 215 | 676 | 235 | 1126 |
| | European | All | 210 | 365 | 203 | 778 |
| | Other | All | 6 | 15 | 14 | 35 |

*Sex refers to the sex of the affected child in a trio.
†A handful of Malays from Singapore are present in the Asian racial/ethnic group of GENEVA. Recall, the GENEVA 'Asian' group consists of various Asian and southeast Asian ethnic groups (subjects recruited from People's Republic of China, Taiwan, South Korea, Singapore, and the Philippines).

Abbreviations: CL, cleft lip; CLP, cleft lip with palate; CP, cleft palate; OFCs, orofacial clefts

**Table S2: Association results for the most significant markers from the 26 loci for CL/P at a suggestive threshold of $10^{-6}$.** The analysis of CL/P is the same as pooled analysis of CL and CLP subtypes. Results from the genetic overlap analysis of CL & CLP using PLACO at these loci are also provided. Analyses are based on all trios from both POFC and GENEVA for a given cleft subtype. The "No. of trios" columns give the numbers of complete informative case-parent trios as used by gTDT.

| Locus | Nearest gene | rsID | Position (hg19) | Effect allele | CL/P gTDT p-value | CL/P gTDT RR | CL/P No. of trios | CL gTDT p-value | CL gTDT RR | CL No. of trios | CLP gTDT p-value | CLP gTDT RR | CLP No. of trios | CL & CLP PLACO p-value |
|---|---|---|---|---|---|---|---|---|---|---|---|---|---|---|
| 1p36.13 | *PAX7* | rs11308758 | 18959137 | C | $2.8 \times 10^{-11}$ | 1.37 | 1444 | $6.6 \times 10^{-2}$ | 1.18 | 391 | $3.0 \times 10^{-11}$ | 1.45 | 1053 | $1.5 \times 10^{-6}$ |
| 1p22.1 | *ABCA4/ ARHGAP29* | rs560426 | 94553438 | C | $3.2 \times 10^{-13}$ | 1.33 | 2030 | $2.1 \times 10^{-4}$ | 1.34 | 508 | $3.6 \times 10^{-10}$ | 1.33 | 1522 | $2.4 \times 10^{-11}$ |
| 1q32.2 | *IRF6* | rs11119346 | 209983900 | T | $2.0 \times 10^{-21}$ | 0.60 | 1187 | $8.1 \times 10^{-8}$ | 0.54 | 269 | $3.2 \times 10^{-15}$ | 0.62 | 918 | $1.5 \times 10^{-19}$ |
| 2p25.1 | *AC007463.2* | rs35740468 | 8038388 | T | $7.7 \times 10^{-7}$ | 0.79 | 1366 | $6.6 \times 10^{-3}$ | 0.78 | 366 | $3.7 \times 10^{-5}$ | 0.79 | 1000 | $4.2 \times 10^{-6}$ |
| 2p24.2 | *FAM49A* | rs4608519 | 16722908 | G | $8.6 \times 10^{-9}$ | 0.79 | 1874 | $7.1 \times 10^{-2}$ | 0.86 | 479 | $4.0 \times 10^{-7}$ | 0.79 | 1395 | $3.4 \times 10^{-5}$ |
| 3p11.1 | *EPHA3* | rs11918555 | 89541934 | T | $6.9 \times 10^{-9}$ | 0.77 | 1636 | $3.8 \times 10^{-3}$ | 0.78 | 421 | $5.2 \times 10^{-7}$ | 0.77 | 1215 | $1.5 \times 10^{-7}$ |
| 3q12.1 | *COL8A1* | rs793488 | 99495567 | T | $3.2 \times 10^{-9}$ | 0.75 | 1420 | $1.2 \times 10^{-5}$ | 0.64 | 353 | $1.7 \times 10^{-5}$ | 0.78 | 1067 | $1.9 \times 10^{-9}$ |
| 3q26.31 | *NAALADL2* | rs144885328 | 174719127 | G | $3.4 \times 10^{-7}$ | 1.42 | 808 | $7.6 \times 10^{-4}$ | 1.62 | 190 | $7.8 \times 10^{-5}$ | 1.36 | 618 | $5.0 \times 10^{-7}$ |
| 3q28 | *TP63* | rs74914009 | 189545021 | G | $1.7 \times 10^{-7}$ | 1.57 | 539 | $5.3 \times 10^{-4}$ | 1.79 | 144 | $6.1 \times 10^{-5}$ | 1.49 | 395 | $2.8 \times 10^{-7}$ |
| 4p13 | *LIMCH1* | rs28609344 | 41640131 | T | $4.1 \times 10^{-7}$ | 0.77 | 1269 | $7.3 \times 10^{-3}$ | 0.77 | 337 | $1.8 \times 10^{-5}$ | 0.77 | 932 | $3.1 \times 10^{-6}$ |
| 4q21.1 | *SHROOM3* | rs17002103 | 77512356 | G | $5.8 \times 10^{-7}$ | 1.23 | 1839 | $5.6 \times 10^{-2}$ | 1.17 | 460 | $3.1 \times 10^{-6}$ | 1.25 | 1379 | $4.4 \times 10^{-5}$ |
| 6q22.31 | *CLVS2* | rs9490655 | 123417960 | A | $5.0 \times 10^{-7}$ | 0.81 | 1727 | $7.3 \times 10^{-1}$ | 1.03 | 416 | $8.7 \times 10^{-1}$ | 0.99 | 1311 | $8.6 \times 10^{-1}$ |
| 8p12 | *NRG1* | rs1878918 | 32333570 | C | $6.2 \times 10^{-7}$ | 1.23 | 1869 | $2.0 \times 10^{-2}$ | 1.21 | 461 | $1.1 \times 10^{-5}$ | 1.23 | 1408 | $1.1 \times 10^{-5}$ |
| 8q21.3 | *DCAF4L2* | rs12543318 | 88868340 | C | $3.3 \times 10^{-12}$ | 1.31 | 2032 | $4.5 \times 10^{-5}$ | 1.38 | 509 | $1.3 \times 10^{-8}$ | 1.29 | 1523 | $2.5 \times 10^{-11}$ |
| 8q24.21 | 8q24 | rs17242358 | 129964873 | A | $1.4 \times 10^{-33}$ | 2.08 | 1028 | $5.4 \times 10^{-10}$ | 2.06 | 279 | $3.9 \times 10^{-25}$ | 2.09 | 749 | $5.8 \times 10^{-29}$ |
| 9q22.2 | *GADD45G* | rs10908902 | 92224825 | A | $5.9 \times 10^{-7}$ | 1.41 | 732 | $1.7 \times 10^{-3}$ | 1.49 | 208 | $9.5 \times 10^{-5}$ | 1.37 | 524 | $1.5 \times 10^{-6}$ |
| 10q24.32 | *BTRC* | rs11190939 | 103093456 | A | $6.8 \times 10^{-7}$ | 0.71 | 784 | $9.4 \times 10^{-3}$ | 0.71 | 203 | $2.4 \times 10^{-5}$ | 0.71 | 581 | $5.4 \times 10^{-6}$ |
| 10q25.3 | *SHTN1* | rs10886042 | 118863209 | A | $9.9 \times 10^{-11}$ | 1.32 | 1745 | $1.5 \times 10^{-2}$ | 1.23 | 436 | $1.4 \times 10^{-9}$ | 1.35 | 1309 | $1.2 \times 10^{-7}$ |
| 13q31.1 | *SPRY2* | rs1854110 | 80701485 | C | $3.0 \times 10^{-10}$ | 1.31 | 1717 | $1.7 \times 10^{-1}$ | 1.12 | 441 | $8.9 \times 10^{-11}$ | 1.39 | 1276 | $4.2 \times 10^{-5}$ |
| 13q32.3 | *CLYBL* | rs9513637 | 100252331 | T | $5.4 \times 10^{-7}$ | 1.26 | 1522 | $7.4 \times 10^{-2}$ | 1.17 | 394 | $2.0 \times 10^{-6}$ | 1.29 | 1128 | $6.7 \times 10^{-5}$ |
| 14q22.1 | *GNG2* | rs144433632 | 51856064 | AAT | $5.3 \times 10^{-8}$ | 0.80 | 1812 | $1.4 \times 10^{-3}$ | 0.77 | 464 | $9.2 \times 10^{-6}$ | 0.81 | 1348 | $2.1 \times 10^{-7}$ |
| 17p13.1 | *NTN1* | rs12944377 | 8947708 | C | $1.6 \times 10^{-18}$ | 0.69 | 1809 | $3.5 \times 10^{-4}$ | 0.74 | 447 | $6.8 \times 10^{-16}$ | 0.67 | 1362 | $9.1 \times 10^{-14}$ |
| 17q22 | *C17orf67* | rs227727 | 54776955 | T | $4.6 \times 10^{-8}$ | 1.24 | 1991 | $8.6 \times 10^{-4}$ | 1.30 | 501 | $1.2 \times 10^{-5}$ | 1.22 | 1490 | $1.4 \times 10^{-7}$ |
| 19q13.11 | *RHPN2* | rs10417111 | 33504997 | T | $7.9 \times 10^{-8}$ | 0.66 | 654 | $5.2 \times 10^{-1}$ | 0.91 | 166 | $6.0 \times 10^{-9}$ | 0.60 | 488 | $9.7 \times 10^{-3}$ |
| 20q12 | *MAFB* | rs11698990 | 39271008 | G | $7.2 \times 10^{-11}$ | 0.78 | 2042 | $2.4 \times 10^{-2}$ | 0.84 | 503 | $5.1 \times 10^{-10}$ | 0.76 | 1539 | $2.4 \times 10^{-7}$ |
| 21q11.2 | *PPP6R2P1* | rs1297095 | 15380777 | T | $8.9 \times 10^{-7}$ | 1.25 | 1601 | $3.3 \times 10^{-3}$ | 1.29 | 416 | $7.3 \times 10^{-5}$ | 1.23 | 1185 | $2.6 \times 10^{-6}$ |

Abbreviations: Chr, chromosome; CL, cleft lip; CLP, cleft lip with palate; CL/P, cleft lip with or without palate; gTDT, genotypic transmission disequilibrium test; PLACO, pleiotropic analysis under composite null hypothesis; RR, relative risk (with respect to the reported effect allele)

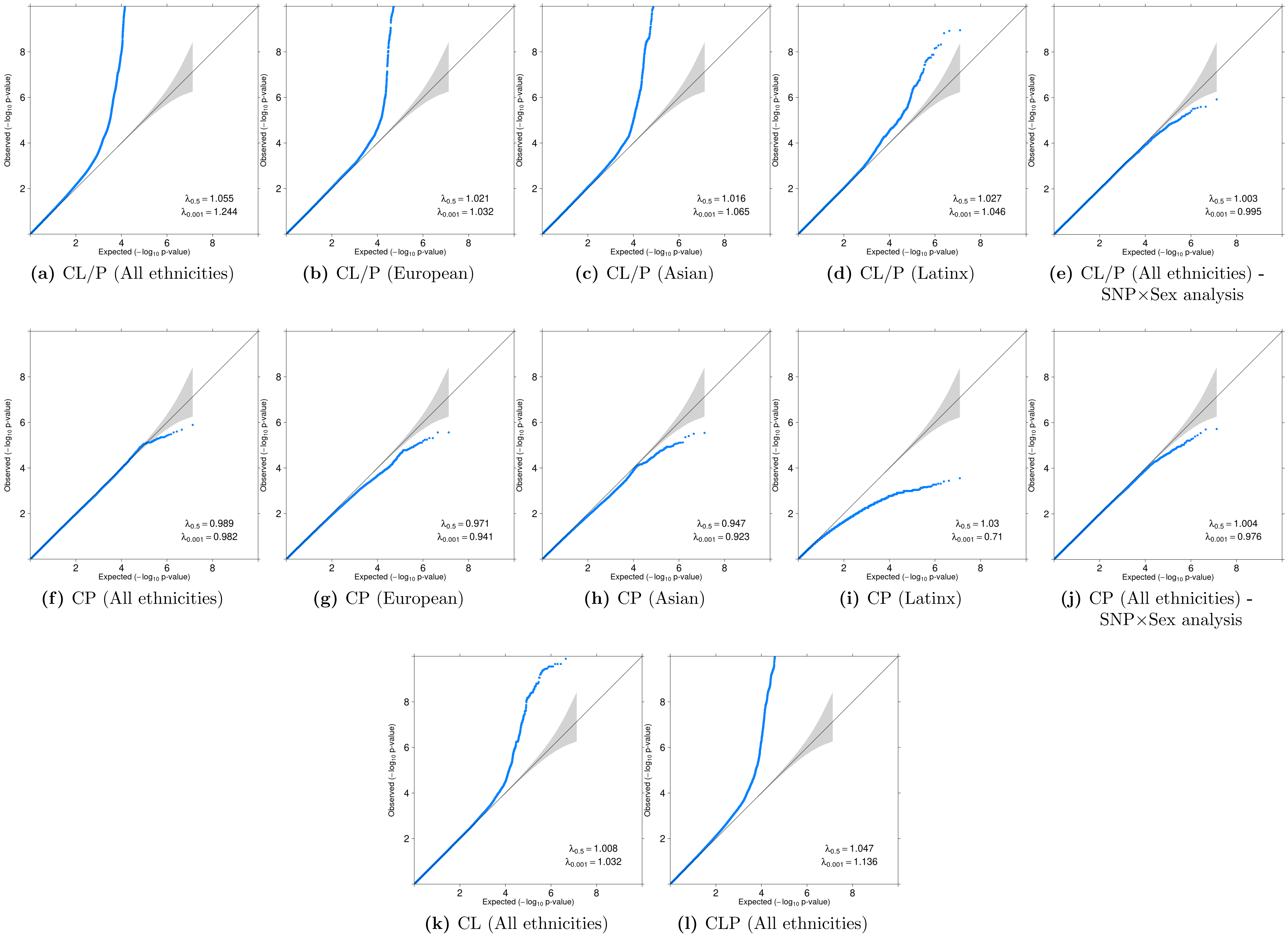


**(a)** CL/P (All ethnicities) **(b)** CL/P (European) **(c)** CL/P (Asian) **(d)** CL/P (Latinx) **(e)** CL/P (All ethnicities) - SNP×Sex analysis

**(f)** CP (All ethnicities) **(g)** CP (European) **(h)** CP (Asian) **(i)** CP (Latinx) **(j)** CP (All ethnicities) - SNP×Sex analysis

**(k)** CL (All ethnicities) **(l)** CLP (All ethnicities)

**Figure S1: QQ plots of the gTDT summary statistics used for different genetic overlap analysis using PLACO in this manuscript.**

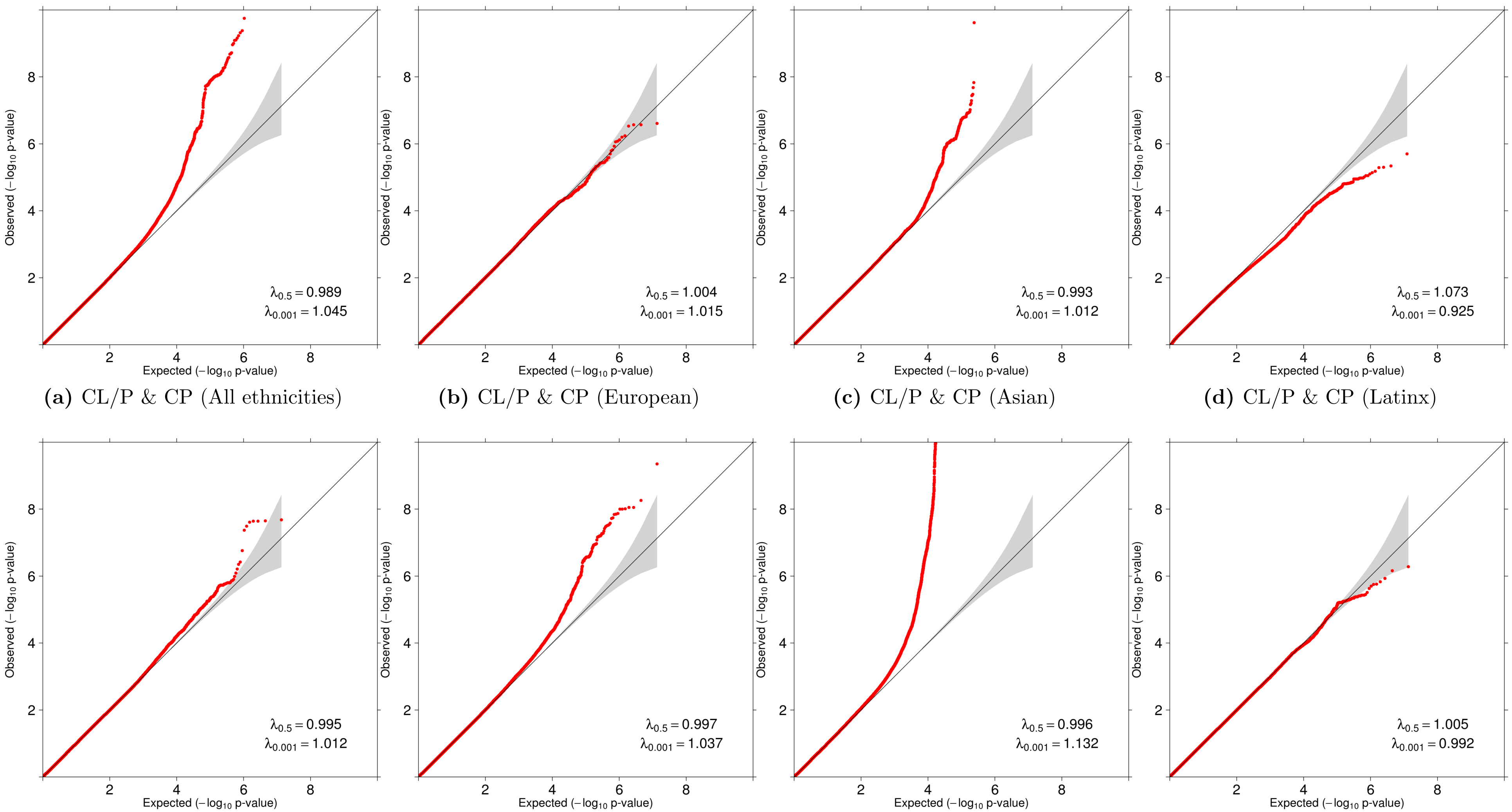


**(a)** CL/P & CP (All ethnicities) **(b)** CL/P & CP (European) **(c)** CL/P & CP (Asian) **(d)** CL/P & CP (Latinx)

**(e)** CL & CP (All ethnicities) **(f)** CLP & CP (All ethnicities) **(g)** CL & CLP (All ethnicities) **(h)** CL/P & CP (All ethnicities) - SNP×Sex analysis

**Figure S2: QQ plots from the different PLACO analyses, including stratified analyses, conducted in this manuscript.**

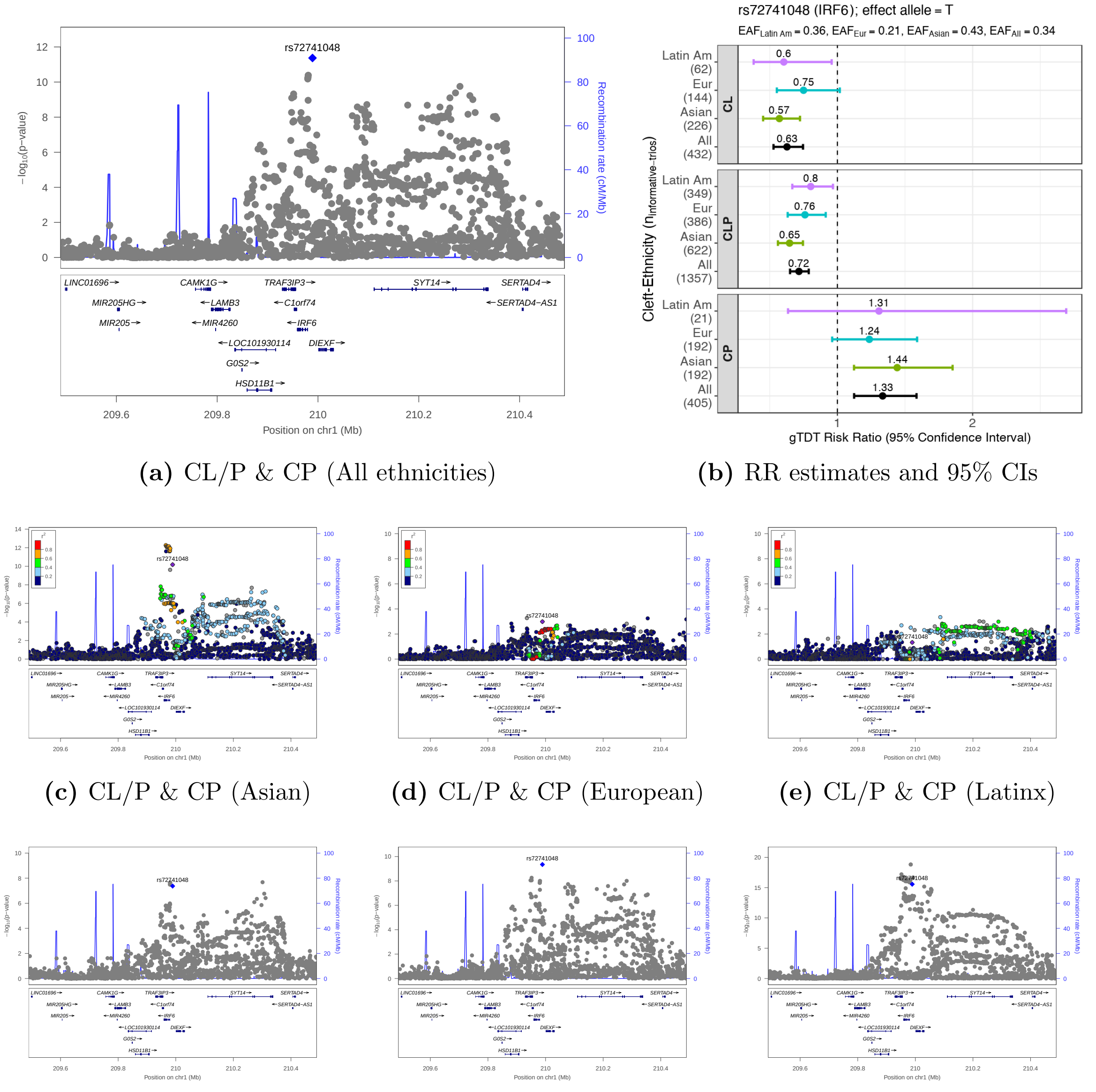


**(a)** CL/P & CP (All ethnicities) **(b)** RR estimates and 95% CIs

**(c)** CL/P & CP (Asian) **(d)** CL/P & CP (European) **(e)** CL/P & CP (Latinx)

**(f)** CL & CP (All ethnicities) **(g)** CLP & CP (All ethnicities) **(h)** CL & CLP (All ethnicities)

**Figure S3: Regional association plots for 1q32.2 (*IRF6*) identified as a region of genetic overlap between CL/P & CP.** LocusZoom plots focus on PLACO analysis of **(a)** CL/P & CP, **(c)** CL/P & CP in Asian ancestry, **(d)** CL/P & CP in European ancestry, **(e)** CL/P & CP in Latin American ancestry, **(f)** CL & CP, **(g)** CLP & CP, **(h)** CL & CLP. The blue or purple diamond represents the most strongly associated SNP in the region showing evidence of genetic overlap. For stratified analyses across racial/ethnic groups, the colors of the SNPs represent their LD with the most strongly associated SNP, as shown in the color legend. For combined multi-ethnic analyses, there is no unique LD between SNPs and hence no color has been used. Panel **(b)** shows relative risk estimates and their 95% confidence intervals as obtained from the gTDT analyses.

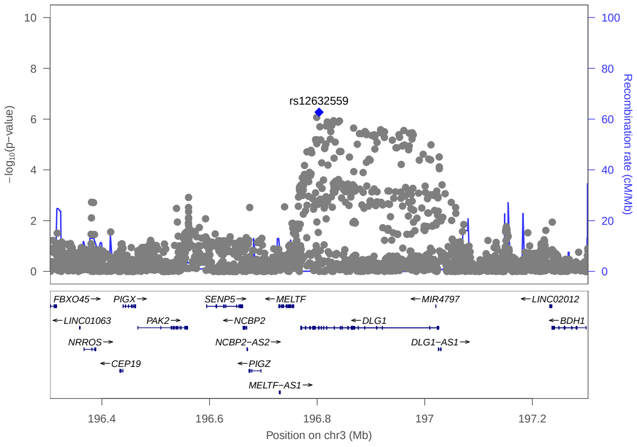


**(a)** CL/P & CP (All ethnicities)

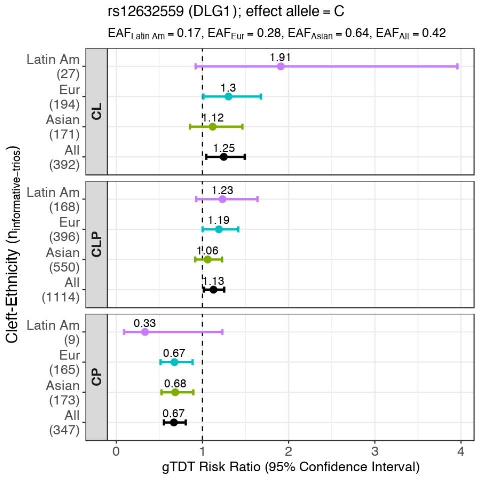


**(b)** RR estimates and 95% CIs

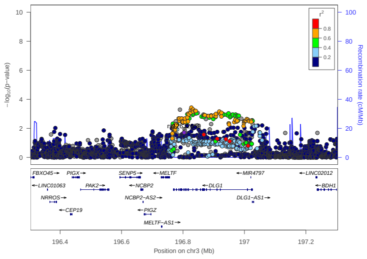

**(c)** CL/P & CP (Asian)

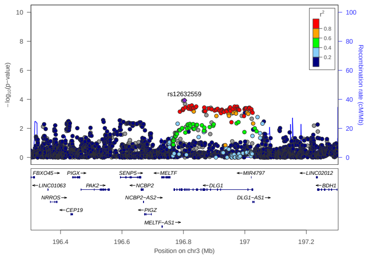

**(d)** CL/P & CP (European)

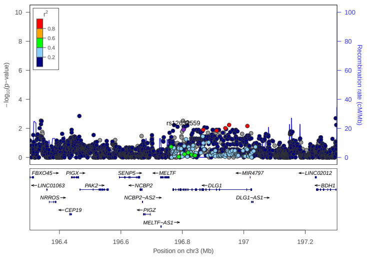

**(e)** CL/P & CP (Latinx)

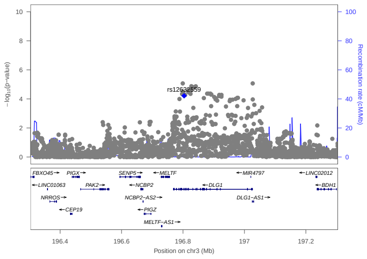

**(f)** CL & CP (All ethnicities)

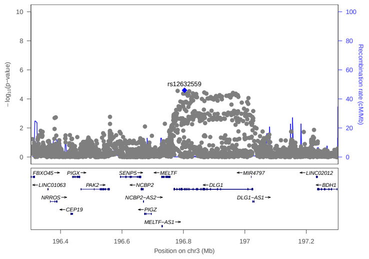

**(g)** CLP & CP (All ethnicities)

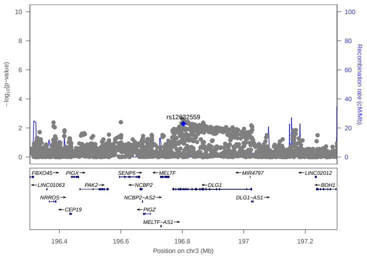

**(h)** CL & CLP (All ethnicities)

**Figure S4: Regional association plots for 3q29 (*DLG1*) identified as a region of genetic overlap between CL/P & CP.** LocusZoom plots focus on PLACO analysis of **(a)** CL/P & CP, **(c)** CL/P & CP in Asian ancestry, **(d)** CL/P & CP in European ancestry, **(e)** CL/P & CP in Latin American ancestry, **(f)** CL & CP, **(g)** CLP & CP, **(h)** CL & CLP. The blue or purple diamond represents the most strongly associated SNP in the region showing evidence of genetic overlap. For stratified analyses across racial/ethnic groups, the colors of the SNPs represent their LD with the most strongly associated SNP, as shown in the color legend. For combined multi-ethnic analyses, there is no unique LD between SNPs and hence no color has been used. Panel **(b)** shows relative risk estimates and their 95% confidence intervals as obtained from the gTDT analyses.

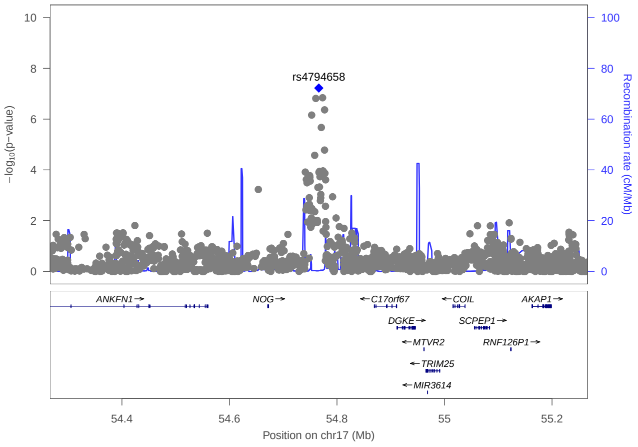


**(a)** CL/P & CP (All ethnicities)

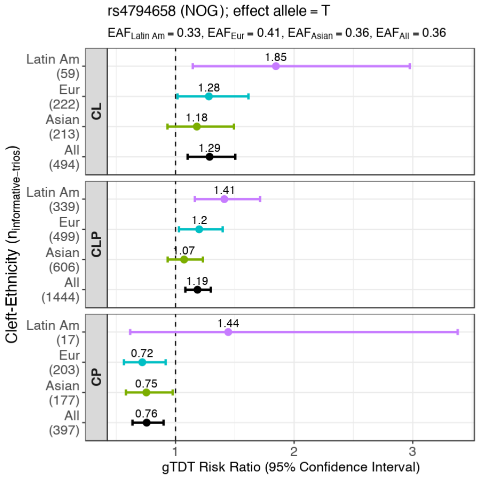


**(b)** RR estimates and 95% CIs

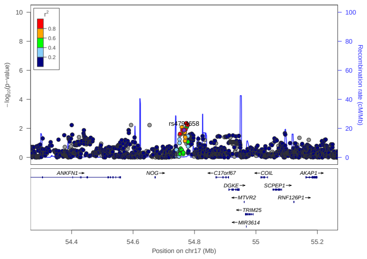

**(c)** CL/P & CP (Asian)

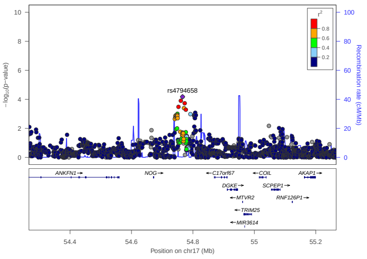

**(d)** CL/P & CP (European)

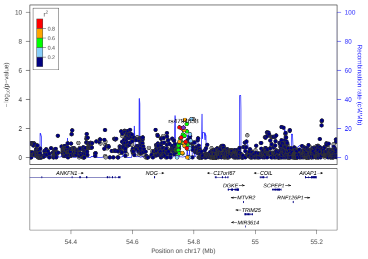

**(e)** CL/P & CP (Latinx)

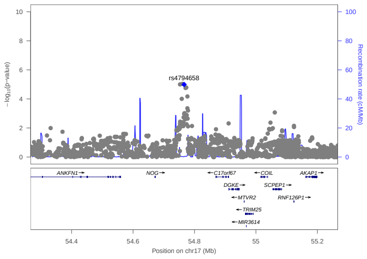

**(f)** CL & CP (All ethnicities)

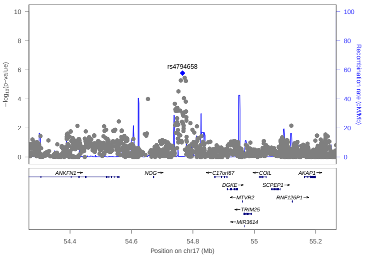

**(g)** CLP & CP (All ethnicities)

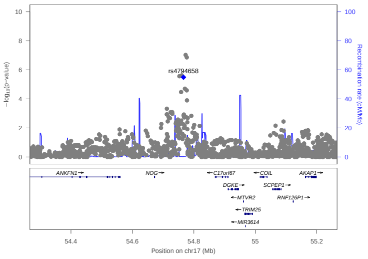

**(h)** CL & CLP (All ethnicities)

**Figure S5: Regional association plots for 17q22 (*NOG*) identified as a region of genetic overlap between CL/P & CP.** LocusZoom plots focus on PLACO analysis of **(a)** CL/P & CP, **(c)** CL/P & CP in Asian ancestry, **(d)** CL/P & CP in European ancestry, **(e)** CL/P & CP in Latin American ancestry, **(f)** CL & CP, **(g)** CLP & CP, **(h)** CL & CLP. The blue or purple diamond represents the most strongly associated SNP in the region showing evidence of genetic overlap. For stratified analyses across racial/ethnic groups, the colors of the SNPs represent their LD with the most strongly associated SNP, as shown in the color legend. For combined multi-ethnic analyses, there is no unique LD between SNPs and hence no color has been used. Panel **(b)** shows relative risk estimates and their 95% confidence intervals as obtained from the gTDT analyses.

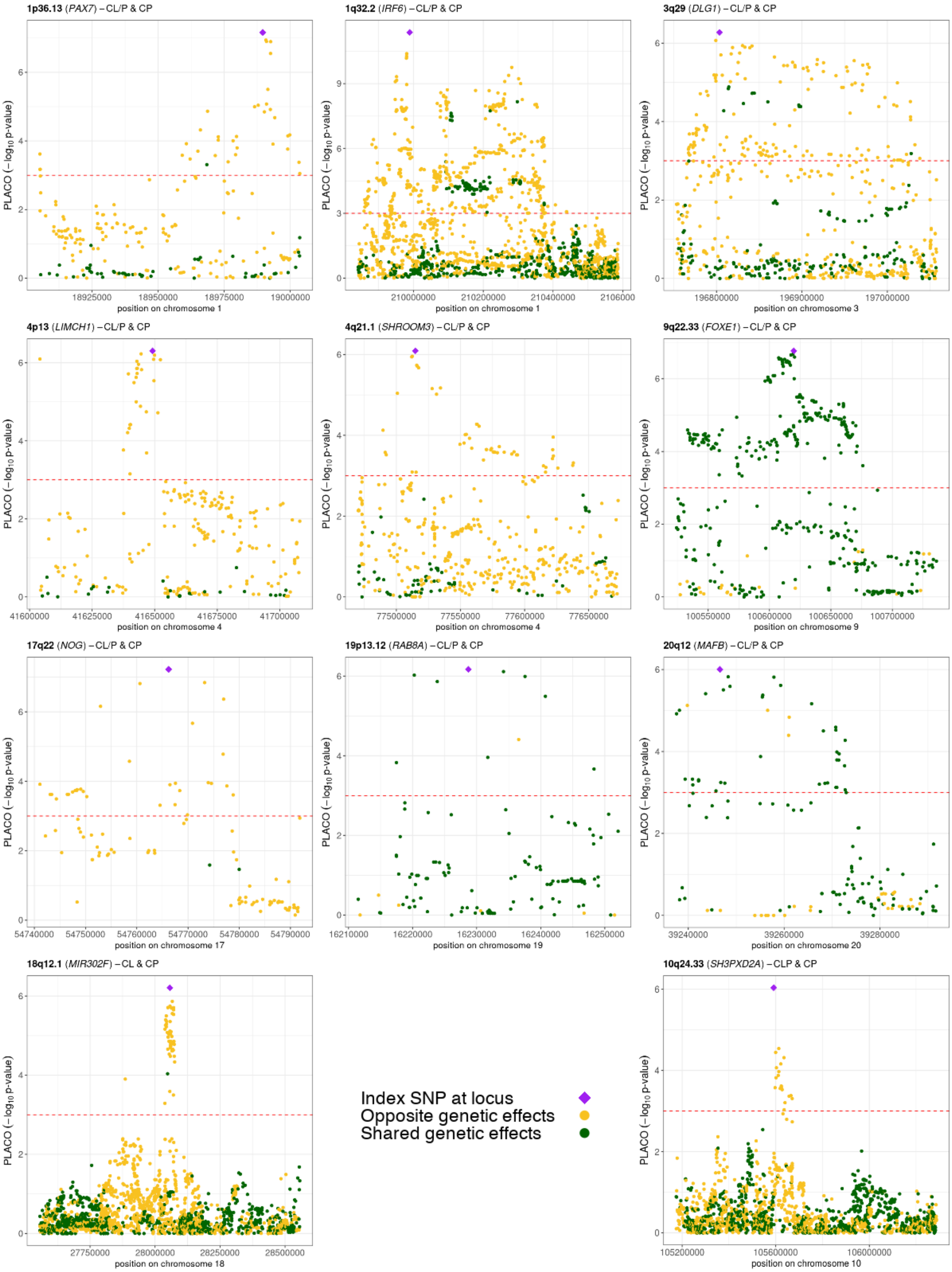


**Figure S6: Regional association plots of PLACO p-values, annotated by directions of effect sizes, for variants in the 9 loci showing statistical evidence of genetic overlap between CL/P & CP, along with 2 additional loci of genetic overlap between component OFC subtypes.** Index SNP here is the lead (most significant) SNP at each locus. SNPs with opposite genetic effects for 2 OFC subgroups are colored in golden yellow while those with shared effects are in dark green. The directions of genetic effects are determined from the relative risk (RR) estimates for each subgroup as provided by the gTDT method. RR estimates and the corresponding 95% confidence intervals for the SNPs above the dashed red horizontal line are portrayed in Figure 2.

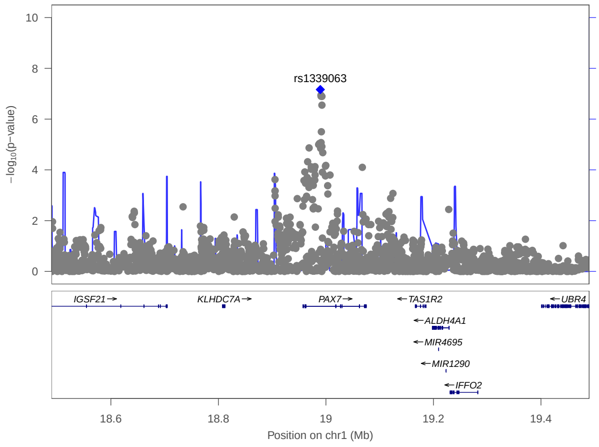


**(a)** CL/P & CP (All ethnicities)

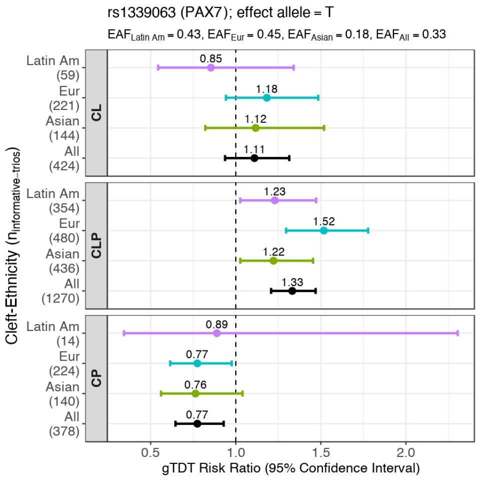


**(b)** RR estimates and 95% CIs

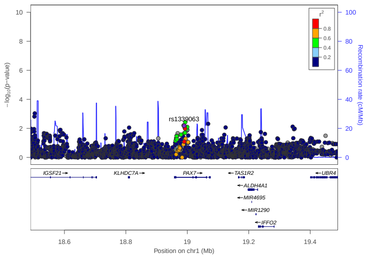

**(c)** CL/P & CP (Asian)

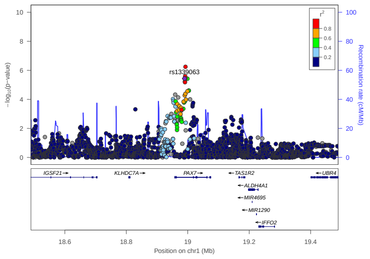

**(d)** CL/P & CP (European)

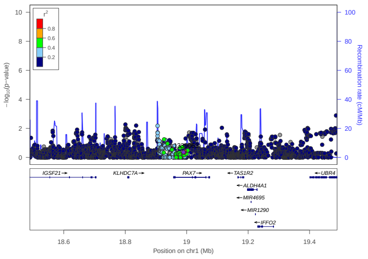

**(e)** CL/P & CP (Latinx)

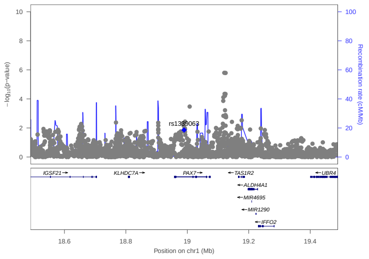

**(f)** CL & CP (All ethnicities)

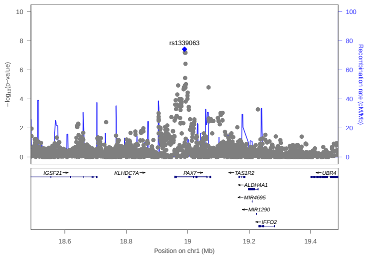

**(g)** CLP & CP (All ethnicities)

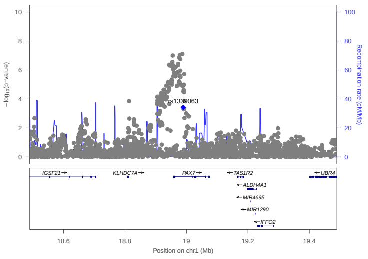

**(h)** CL & CLP (All ethnicities)

**Figure S7: Regional association plots for 1p36.13 (*PAX7*) identified as a region of genetic overlap between CL/P & CP.** LocusZoom plots focus on PLACO analysis of **(a)** CL/P & CP, **(c)** CL/P & CP in Asian ancestry, **(d)** CL/P & CP in European ancestry, **(e)** CL/P & CP in Latin American ancestry, **(f)** CL & CP, **(g)** CLP & CP, **(h)** CL & CLP. The blue or purple diamond represents the most strongly associated SNP in the region showing evidence of genetic overlap. For stratified analyses across racial/ethnic groups, the colors of the SNPs represent their LD with the most strongly associated SNP, as shown in the color legend. For combined multi-ethnic analyses, there is no unique LD between SNPs and hence no color has been used. Panel **(b)** shows relative risk estimates and their 95% confidence intervals as obtained from the gTDT analyses.

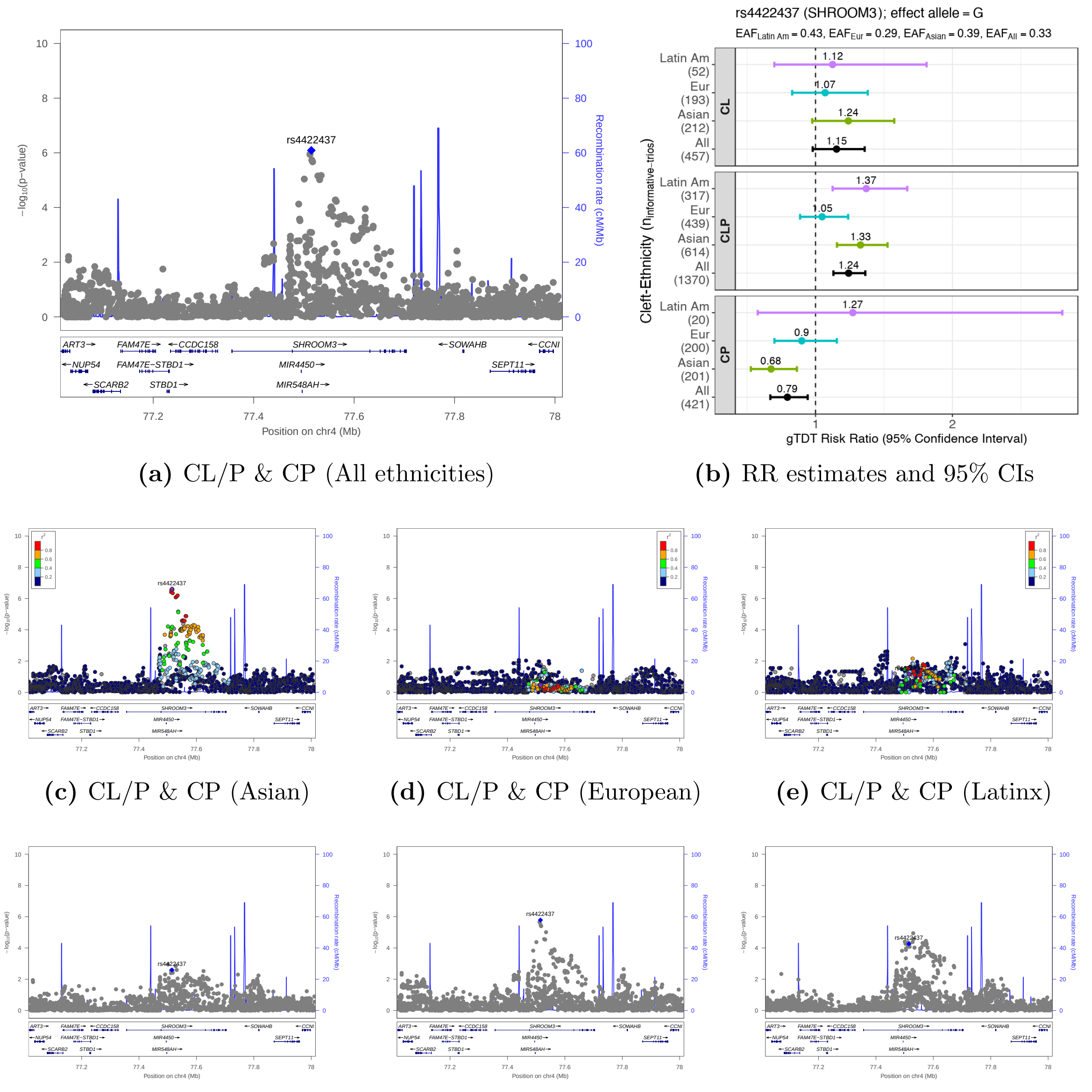


**(a)** CL/P & CP (All ethnicities) **(b)** RR estimates and 95% CIs

**(c)** CL/P & CP (Asian) **(d)** CL/P & CP (European) **(e)** CL/P & CP (Latinx)

**(f)** CL & CP (All ethnicities) **(g)** CLP & CP (All ethnicities) **(h)** CL & CLP (All ethnicities)

**Figure S8: Regional association plots for 4q21.1 (*SHROOM3*) identified as a region of genetic overlap between CL/P & CP.** LocusZoom plots focus on PLACO analysis of **(a)** CL/P & CP, **(c)** CL/P & CP in Asian ancestry, **(d)** CL/P & CP in European ancestry, **(e)** CL/P & CP in Latin American ancestry, **(f)** CL & CP, **(g)** CLP & CP, **(h)** CL & CLP. The blue or purple diamond represents the most strongly associated SNP in the region showing evidence of genetic overlap. For stratified analyses across racial/ethnic groups, the colors of the SNPs represent their LD with the most strongly associated SNP, as shown in the color legend. For combined multi-ethnic analyses, there is no unique LD between SNPs and hence no color has been used. Panel **(b)** shows relative risk estimates and their 95% confidence intervals as obtained from the gTDT analyses.

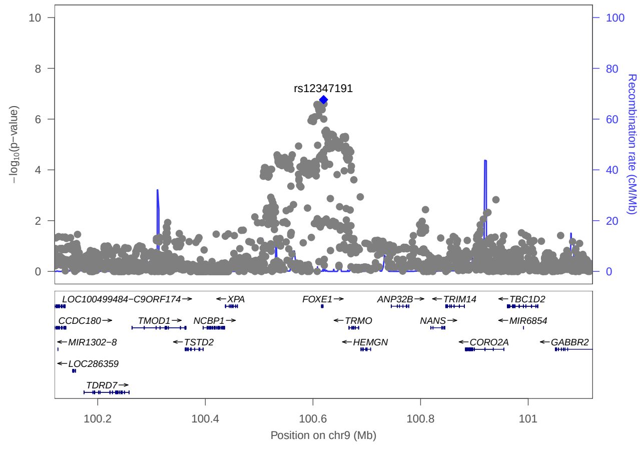


**(a)** CL/P & CP (All ethnicities)

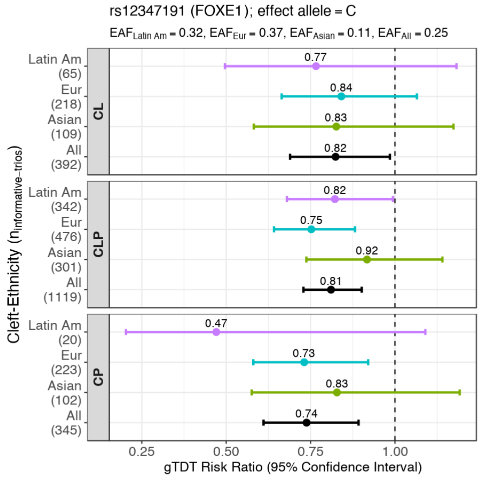


**(b)** RR estimates and 95% CIs

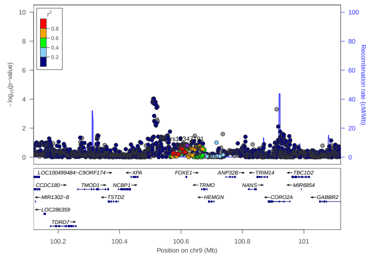

**(c)** CL/P & CP (Asian)

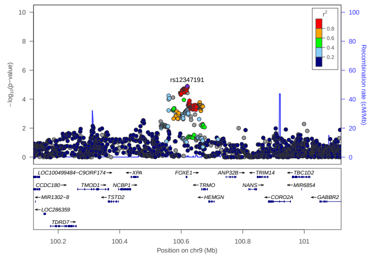


**(d)** CL/P & CP (European)

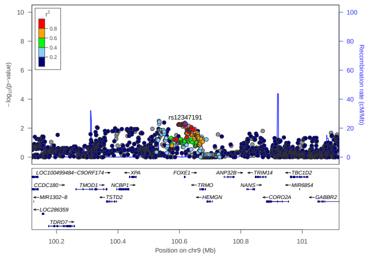


**(e)** CL/P & CP (Latinx)

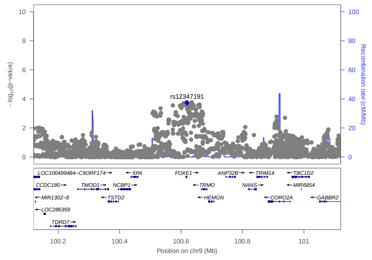


**(f)** CL & CP (All ethnicities)

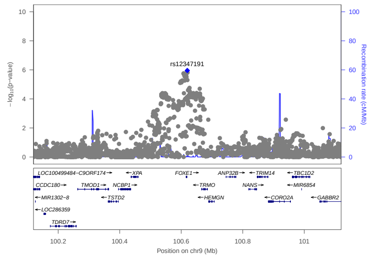


**(g)** CLP & CP (All ethnicities)

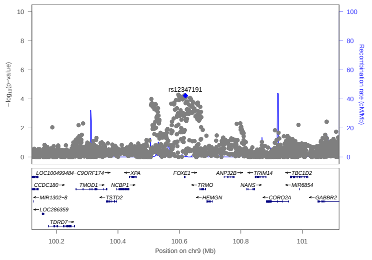


**(h)** CL & CLP (All ethnicities)

**Figure S9: Regional association plots for 9q22.33 (*FOXE1*) identified as a region of genetic overlap between CL/P & CP.** LocusZoom plots focus on PLACO analysis of **(a)** CL/P & CP, **(c)** CL/P & CP in Asian ancestry, **(d)** CL/P & CP in European ancestry, **(e)** CL/P & CP in Latin American ancestry, **(f)** CL & CP, **(g)** CLP & CP, **(h)** CL & CLP. The blue or purple diamond represents the most strongly associated SNP in the region showing evidence of genetic overlap. For stratified analyses across racial/ethnic groups, the colors of the SNPs represent their LD with the most strongly associated SNP, as shown in the color legend. For combined multi-ethnic analyses, there is no unique LD between SNPs and hence no color has been used. Panel **(b)** shows relative risk estimates and their 95% confidence intervals as obtained from the gTDT analyses.

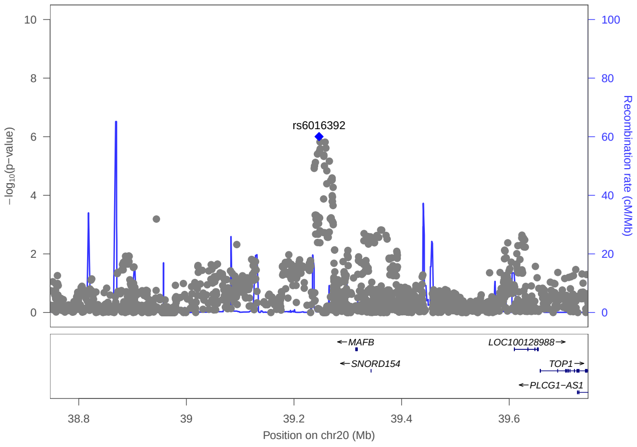


**(a)** CL/P & CP (All ethnicities)

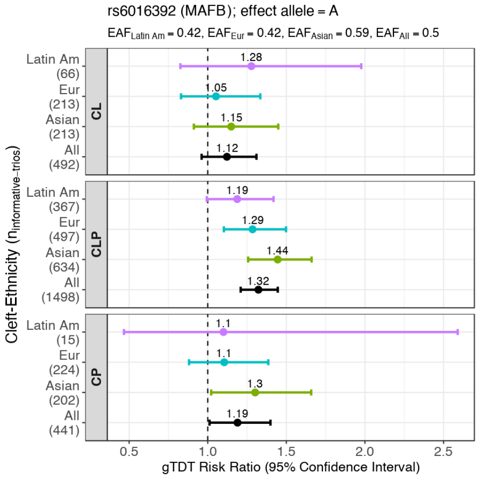


**(b)** RR estimates and 95% CIs

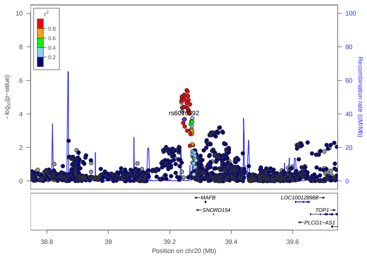

**(c)** CL/P & CP (Asian)

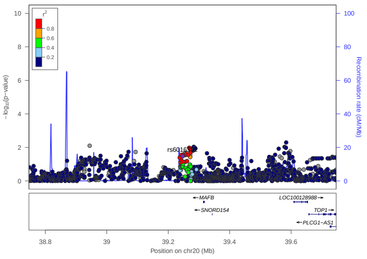

**(d)** CL/P & CP (European)

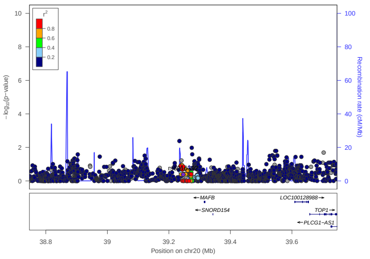

**(e)** CL/P & CP (Latinx)

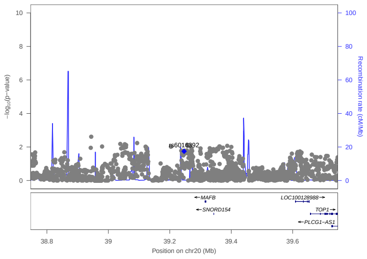

**(f)** CL & CP (All ethnicities)

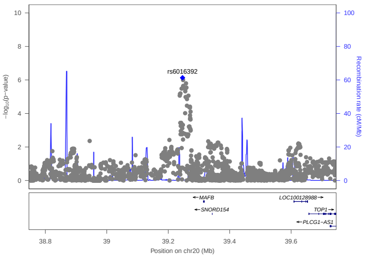

**(g)** CLP & CP (All ethnicities)

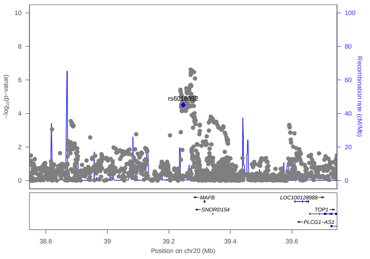

**(h)** CL & CLP (All ethnicities)

**Figure S10: Regional association plots for 20q12 (*MAFB*) identified as a region of genetic overlap between CL/P & CP.** LocusZoom plots focus on PLACO analysis of **(a)** CL/P & CP, **(c)** CL/P & CP in Asian ancestry, **(d)** CL/P & CP in European ancestry, **(e)** CL/P & CP in Latin American ancestry, **(f)** CL & CP, **(g)** CLP & CP, **(h)** CL & CLP. The blue or purple diamond represents the most strongly associated SNP in the region showing evidence of genetic overlap. For stratified analyses across racial/ethnic groups, the colors of the SNPs represent their LD with the most strongly associated SNP, as shown in the color legend. For combined multi-ethnic analyses, there is no unique LD between SNPs and hence no color has been used. Panel **(b)** shows relative risk estimates and their 95% confidence intervals as obtained from the gTDT analyses.

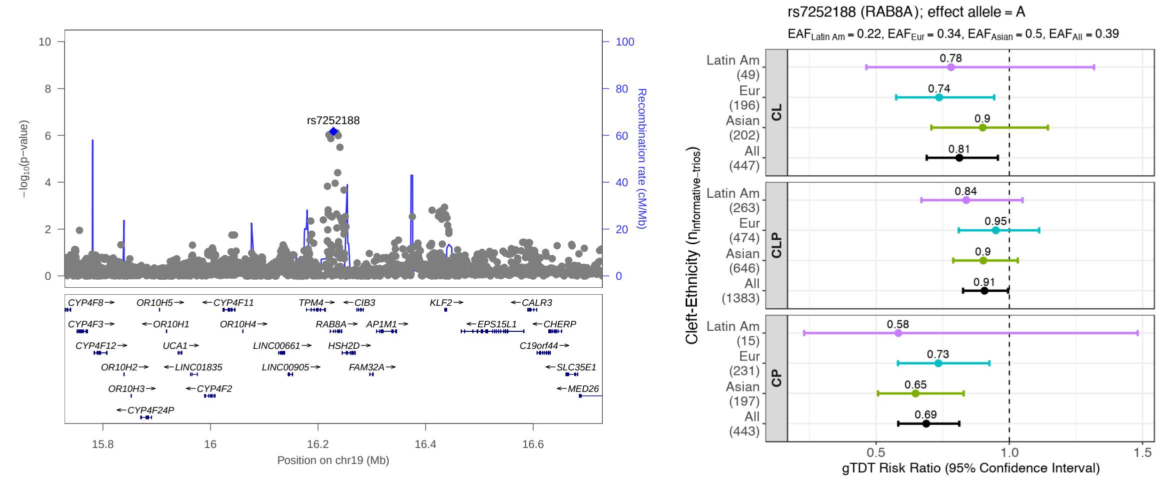


**(a)** CL/P & CP (All ethnicities) **(b)** RR estimates and 95% CIs

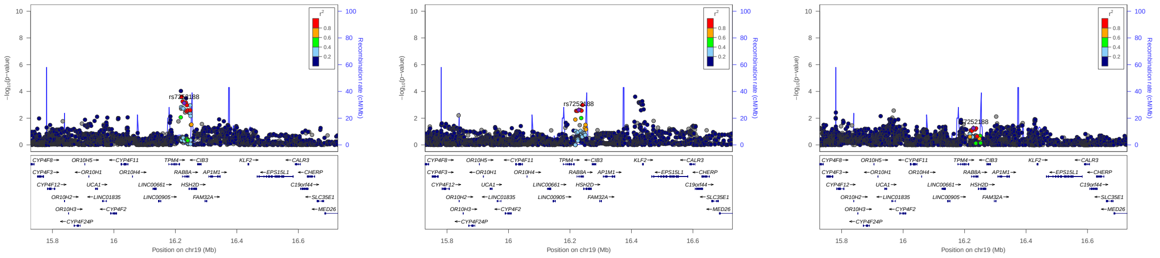

**(c)** CL/P & CP (Asian) **(d)** CL/P & CP (European) **(e)** CL/P & CP (Latinx)

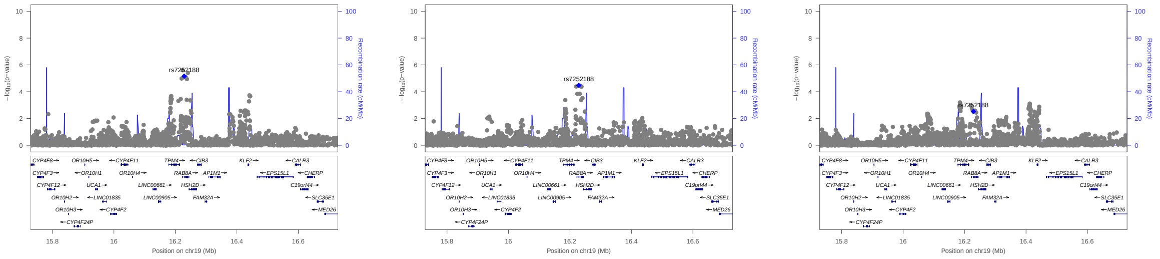

**(f)** CL & CP (All ethnicities) **(g)** CLP & CP (All ethnicities) **(h)** CL & CLP (All ethnicities)

**Figure S11: Regional association plots for 19p13.12 (*RAB8A*) identified as a region of genetic overlap between CL/P & CP.** LocusZoom plots focus on PLACO analysis of **(a)** CL/P & CP, **(c)** CL/P & CP in Asian ancestry, **(d)** CL/P & CP in European ancestry, **(e)** CL/P & CP in Latin American ancestry, **(f)** CL & CP, **(g)** CLP & CP, **(h)** CL & CLP. The blue or purple diamond represents the most strongly associated SNP in the region showing evidence of genetic overlap. For stratified analyses across racial/ethnic groups, the colors of the SNPs represent their LD with the most strongly associated SNP, as shown in the color legend. For combined multi-ethnic analyses, there is no unique LD between SNPs and hence no color has been used. Panel **(b)** shows relative risk estimates and their 95% confidence intervals as obtained from the gTDT analyses.

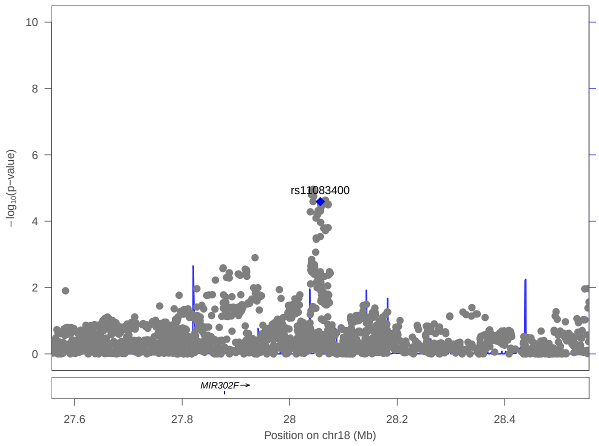


(a) CL/P & CP (All ethnicities)

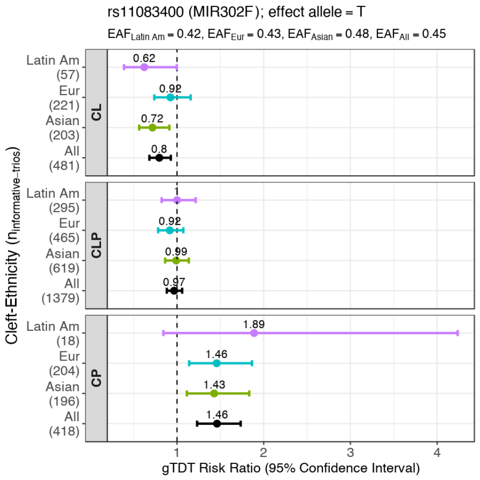


(b) RR estimates and 95% CIs

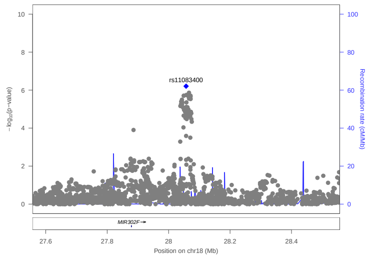


(c) CL & CP (All ethnicities)

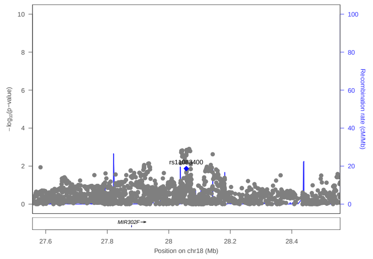


(d) CLP & CP (All ethnicities)

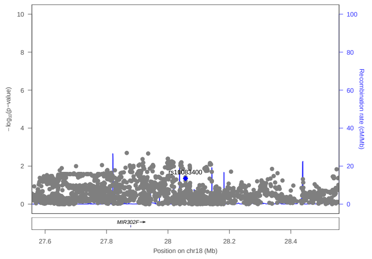


(e) CL & CLP (All ethnicities)

**Figure S12: Regional association plots for 18q12.1 (*MIR302F*) identified as a region of genetic overlap between CL & CP.** LocusZoom plots focus on PLACO analysis of **(a)** CL/P & CP, **(c)** CL & CP, **(d)** CLP & CP, **(e)** CL & CLP. The blue diamond represents the most strongly associated SNP in the region showing evidence of genetic overlap. For multi-ethnic analyses, there is no unique LD between SNPs and hence no color has been used to represent strength of LD. Panel **(b)** shows relative risk estimates and their 95% confidence intervals as obtained from the gTDT analyses.

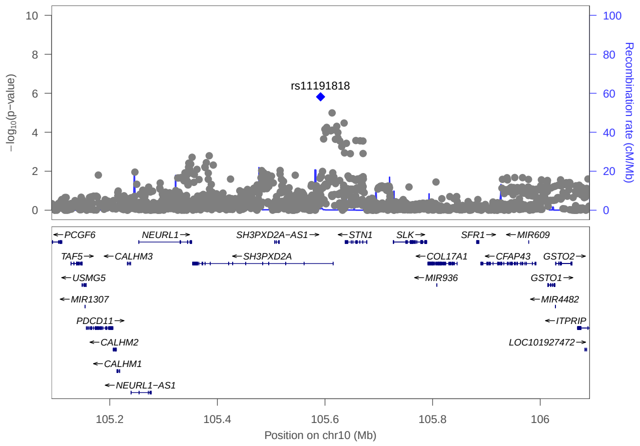


**(a)** CL/P & CP (All ethnicities)

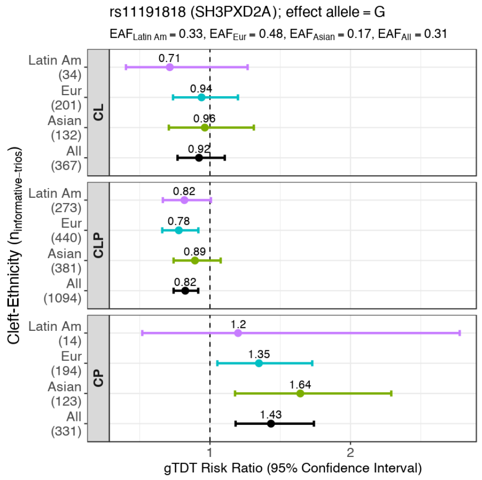


**(b)** RR estimates and 95% CIs

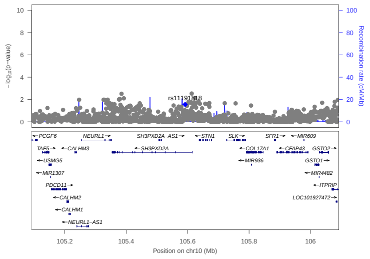

**(c)** CL & CP (All ethnicities)

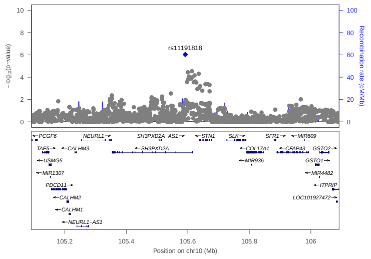


**(d)** CLP & CP (All ethnicities)

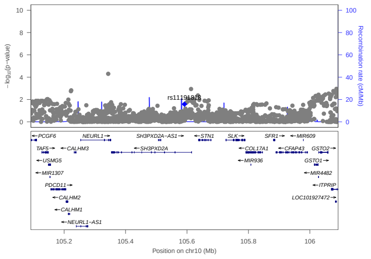

**(e)** CL & CLP (All ethnicities)

**Figure S13: Regional association plots for 10q24.33 (*SH3PXD2A*) identified as a region of genetic overlap between CLP & CP.** LocusZoom plots focus on PLACO analysis of **(a)** CL/P & CP, **(c)** CL & CP, **(d)** CLP & CP, **(e)** CL & CLP. The blue diamond represents the most strongly associated SNP in the region showing evidence of genetic overlap. For multi-ethnic analyses, there is no unique LD between SNPs and hence no color has been used to represent strength of LD. Panel **(b)** shows relative risk estimates and their 95% confidence intervals as obtained from the gTDT analyses.

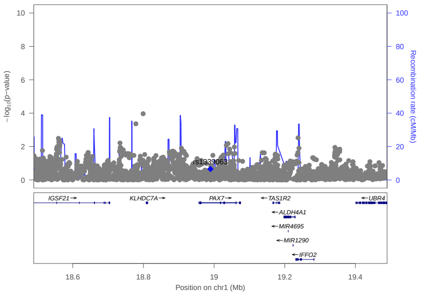

**(a)** *PAX7*

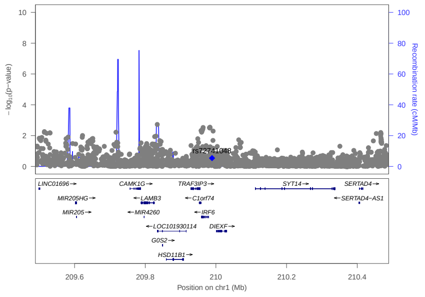

**(b)** *IRF6*

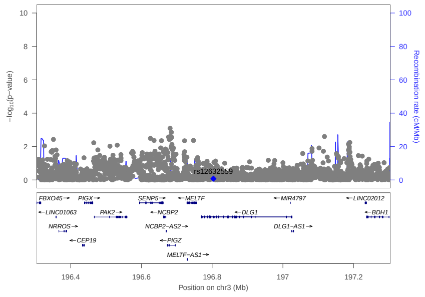

**(c)** *DLG1*

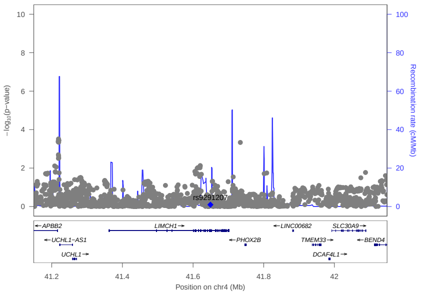

**(d)** *LIMCH1*

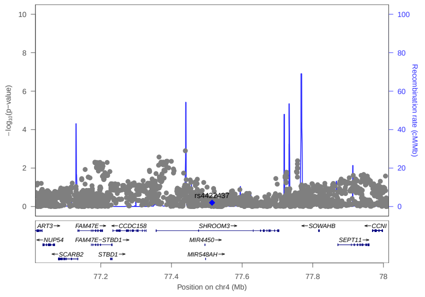

**(e)** *SHROOM3*

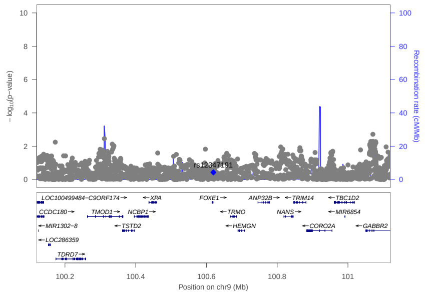

**(f)** *FOXE1*

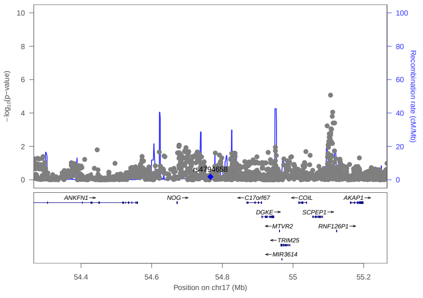

**(g)** *NOG*

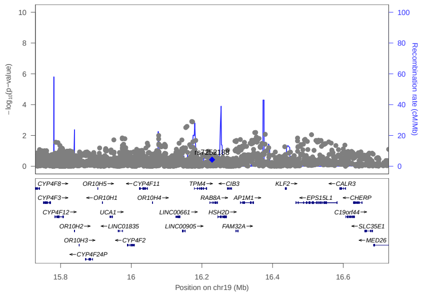

**(h)** *RAB8A*

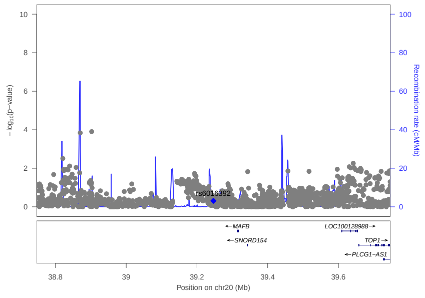

**(i)** *MAFB*

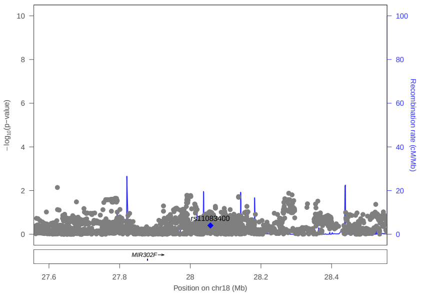

**(j)** *MIR302F*

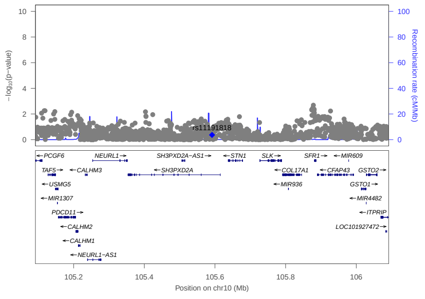

**(k)** *SH3PXD2A*

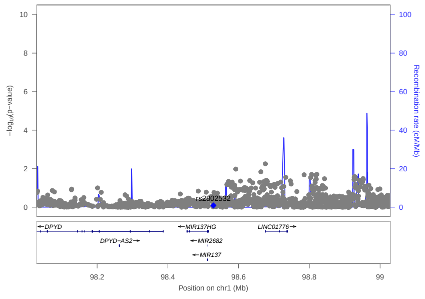

**(l)** *MIR137HG*

**Figure S14: Regional association plots to investigate if genetic overlap between CL/P & CP at any region of interest (as identified by PLACO in different pairwise analyses) is modified by sex.** P-values from our SNP×Sex analyses are plotted. The blue diamond represents the most strongly associated SNP in the region of genetic overlap. For multi-ethnic analyses, there is no unique LD between SNPs and hence no color has been used to represent strength of LD.

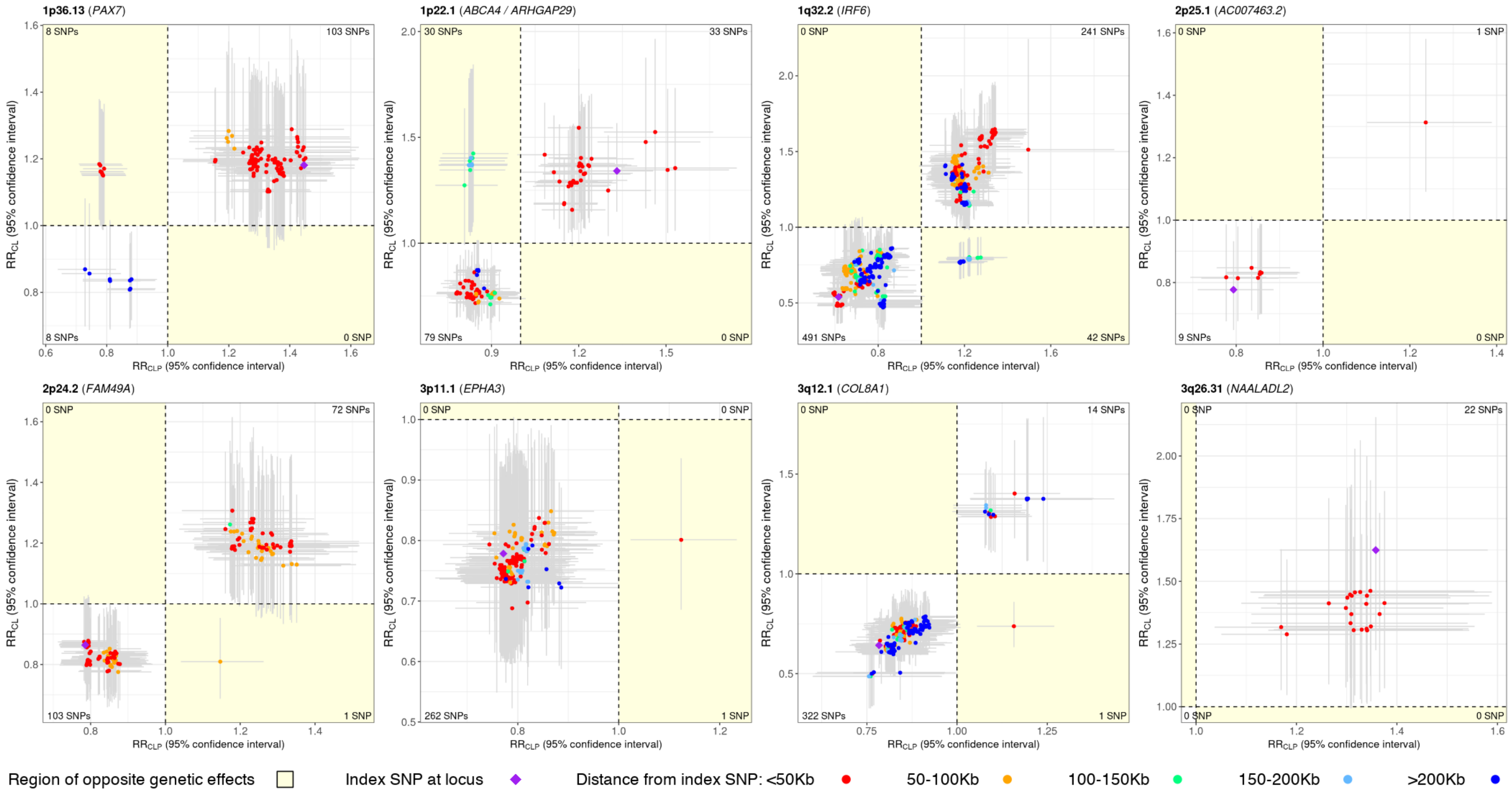


**Figure S15: Scatter plot of relative risk (RR) estimates of CL and CLP, along with corresponding 95% confidence intervals (CIs), for variants in the 26 loci for CL/P.** RR estimates are color annotated based on distance of SNPs from the index/lead SNP. LD-based color annotation is not used since these RR estimates are from multi-ethnic analyses and consequently, there is no unique LD between SNPs. Horizontal (vertical) error bar around each RR estimate corresponds to the 95% CI for the OFC subgroup represented on the x-axis (y-axis). The region depicting opposite genetic effects of SNPs for the 2 OFC subgroups is shaded in yellow. The number of SNPs in each quadrant is printed in the corresponding corner of the plot. While the 26 loci were identified from the gTDT analysis of CL/P at a suggestive threshold of $10^{-6}$, the SNPs plotted here are selected based on two criteria: (i) SNPs in ±500 Kb radius and in LD $r^2 > 0.2$ with the index SNP (the most significant SNP in the locus); and (ii) SNPs with PLACO p-value $< 10^{-3}$ from the genetic overlap analysis of CL & CLP. Two loci, 6q22.31 and 19q13.11 (see Table S2), are not depicted here since no SNP in these 2 loci show significance in PLACO analysis even at a liberal threshold of $10^{-3}$. These plots show the concordance of results for CL/P, and CL & CLP; i.e., the regions of genetic overlap identified by PLACO matches with the shared signals captured by the pooled analysis of CL and CLP subtypes. Additionally, there is some indication of genetically distinct etiology of subtypes CL and CLP at loci 1p36.13 (*PAX7*), 1p22.1 (*ABCA4*, *ARHGAP29*), 1q32.2 (*IRF6*), and 3q12.1 (*COL8A1*) as depicted by the SNPs with opposite genetic effects. **(Figure continues over to next 2 pages)**

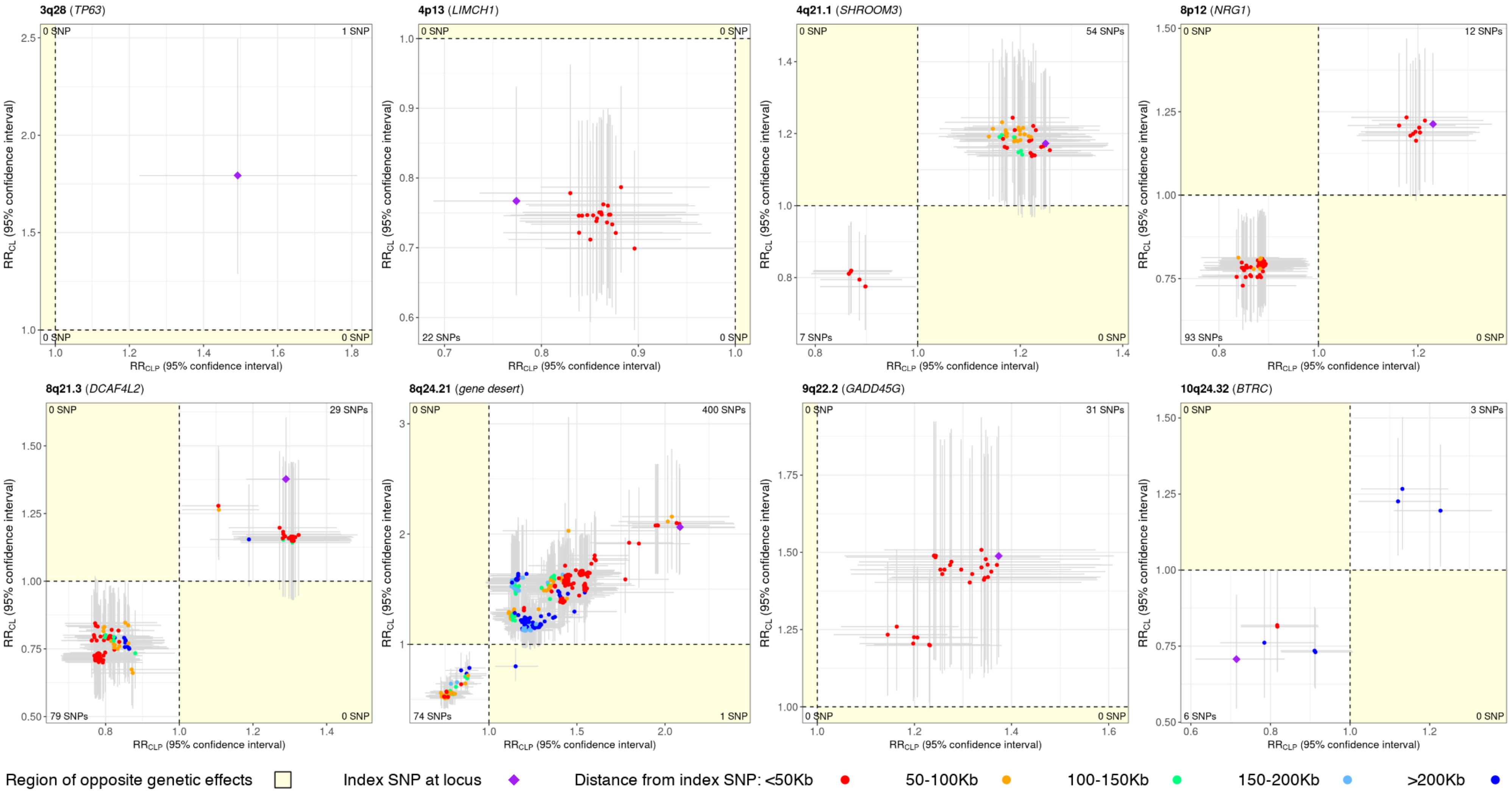


**Figure S15: (Continued from previous page)** See previous page for caption.

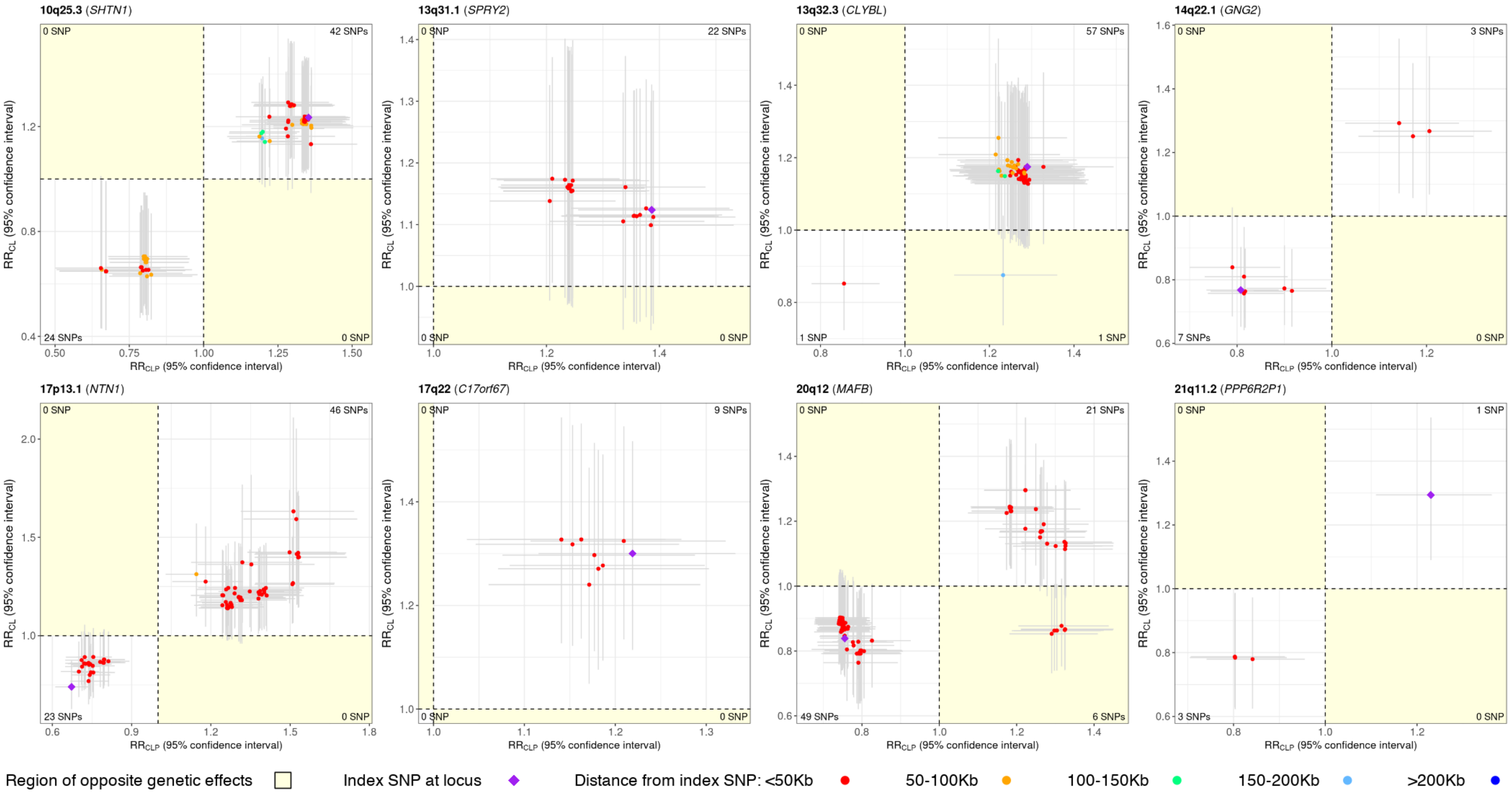


**Figure S15: (Continued from previous page)** See previous page for caption.

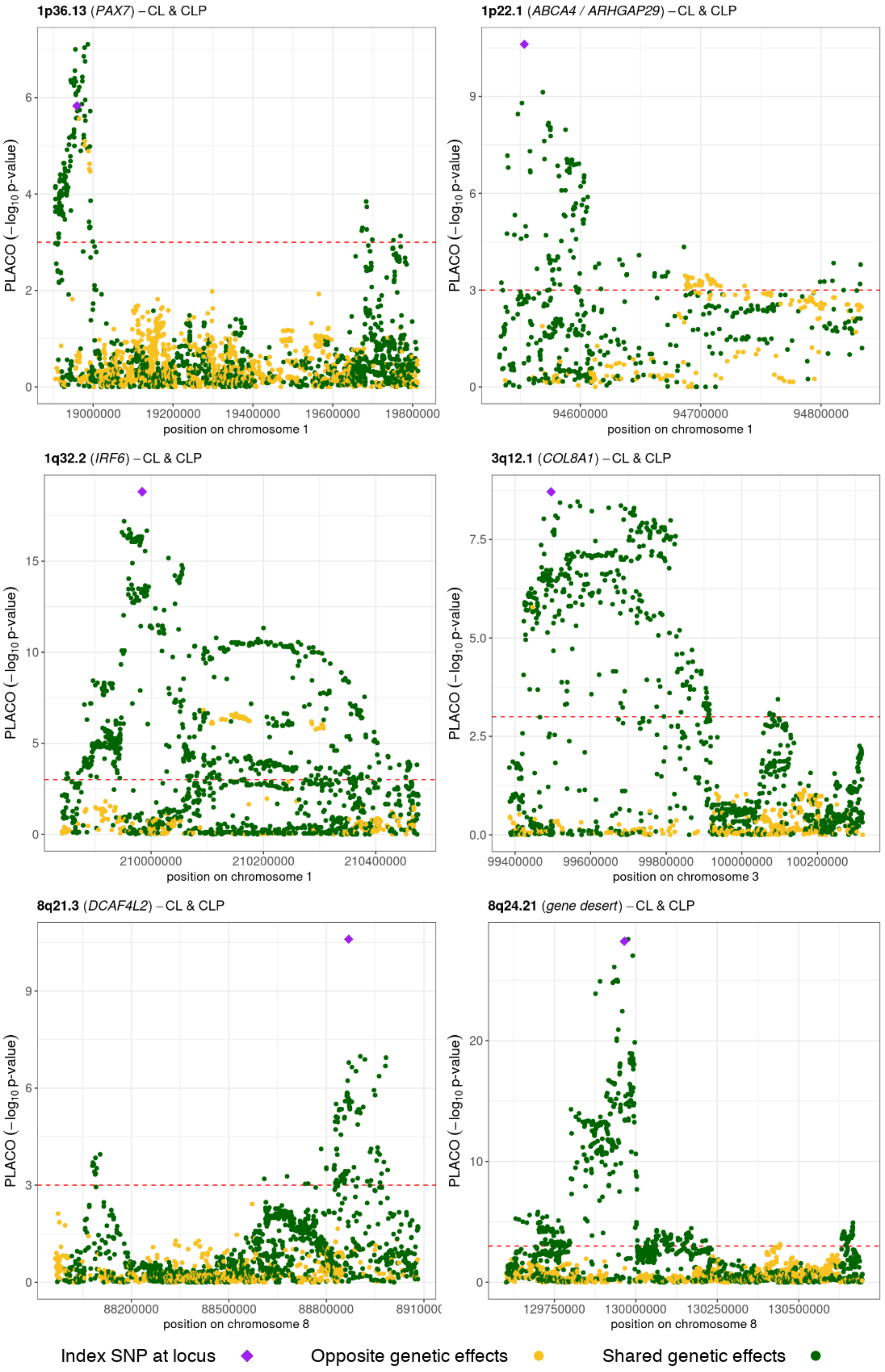


**Figure S16: Regional association plots of PLACO p-values from CL & CLP analysis, annotated by directions of effect sizes, for variants in some of the loci from the 26 loci for CL/P.** Index SNP here is the lead (most significant) SNP from the gTDT analysis of CL/P. SNPs with opposite genetic effects for the 2 OFC subtypes are colored in golden yellow while those with shared effects are in dark green. The directions of genetic effects are determined from the relative risk (RR) estimates for each subtype as provided by the gTDT method. RR estimates and the corresponding 95% confidence intervals for the SNPs above the dashed red horizontal line are portrayed in Figure S15. Each of the loci here appear to have at least 2 distinct regions of genetic overlap.

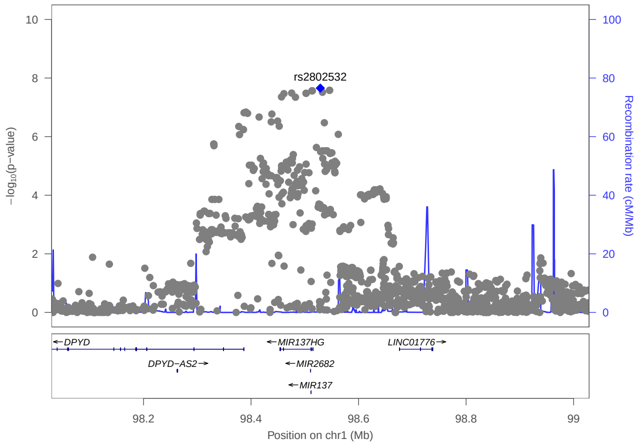


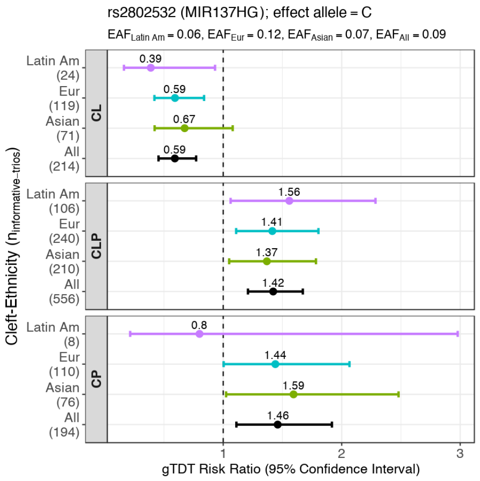


**(a)** CL & CLP (All ethnicities)

**(b)** RR estimates and 95% CIs

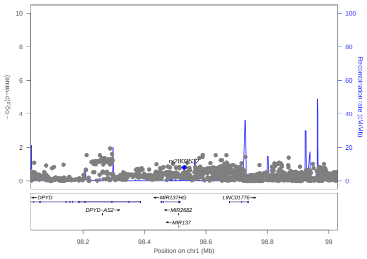

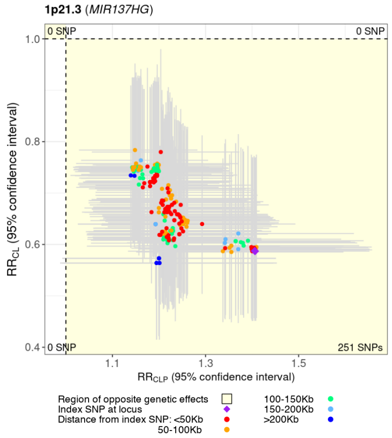


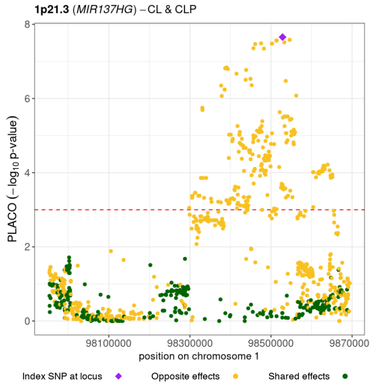


**(c)** Pooled CL + CLP (All ethnicities)

**(d)** Scatter plot of RR estimates – CL & CLP (All ethnicities)

**(e)** CL & CLP (All ethnicities) showing effect directions

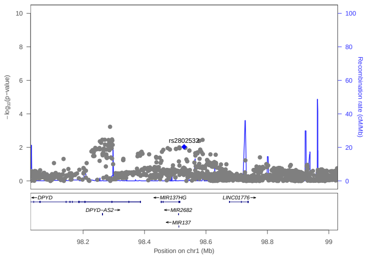

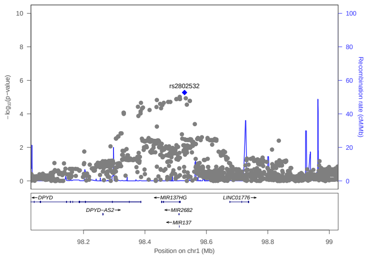

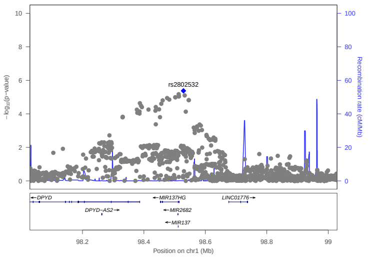

**(f)** CL/P & CP (All ethnicities)

**(g)** CL & CP (All ethnicities)

**(h)** CLP & CP (All ethnicities)

**Figure S17: Regional association plots for 1p21.3 (*MIR137HG*) identified as a region of genetic overlap between CL & CLP.** LocusZoom plots focus on PLACO analysis of **(a)** CL & CLP, **(e)** CL & CLP annoted by directions of genetic effects, **(f)** CL/P & CP, **(g)** CL & CP, **(h)** CLP & CP, while **(c)** shows pooled method analysis of CL and CLP (i.e., gTDT analysis of subgroup CL/P). The blue diamond represents the most strongly associated SNP in the region showing evidence of genetic overlap. For multi-ethnic analyses, there is no unique LD between SNPs and hence no color has been used to represent strength of LD. Panel **(b)** shows relative risk (RR) estimates and their 95% confidence intervals (CIs) across OFC subtypes and racial/ethnic groups as obtained from the gTDT analyses.

Panel **(d)** shows scatter plot of RR estimates of CL and CLP, along with corresponding 95% CIs, for the top several SNPs. SNPs represented here are in ±500 Kb radius and in LD $r^2 > 0.2$ with the index SNP (the most significant SNP in the locus), and with PLACO p-value $< 10^{-3}$ from the genetic overlap analysis of CL & CLP. RR estimates are color annotated based on distance of SNPs from the index/lead SNP. LD-based color annotation is not used since these RR estimates are from multi-ethnic analyses. Horizontal (vertical) error bar around each RR estimate corresponds to the 95% CI for CLP (CL). The region depicting opposite genetic effects of SNPs for the 2 OFC subtypes is shaded in yellow. The number of SNPs in each quadrant is printed in the corresponding corner of the plot.

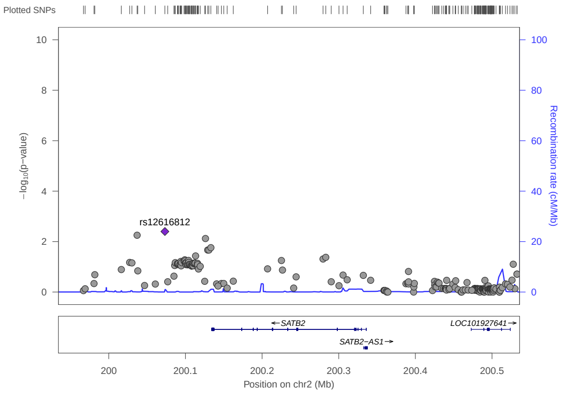


**(a)** *SATB2*

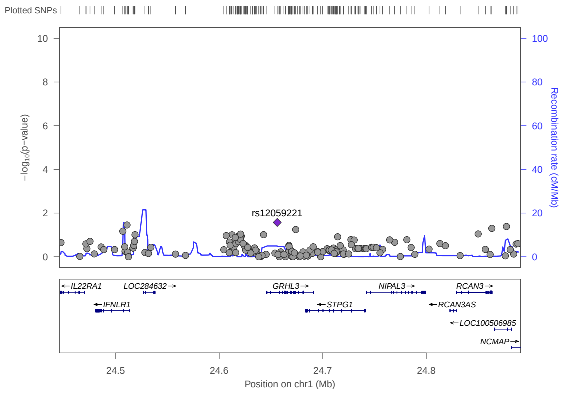


**(b)** *GRHL3*

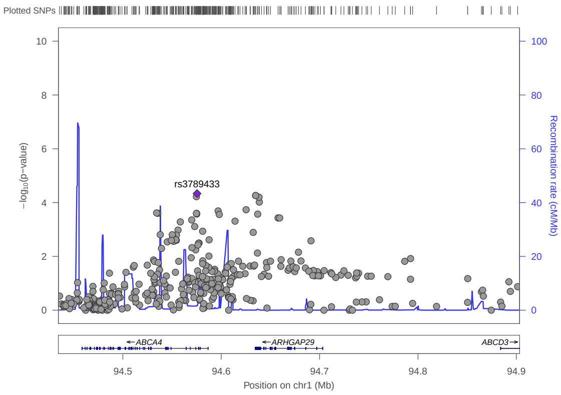


**(c)** *ARHGAP29*

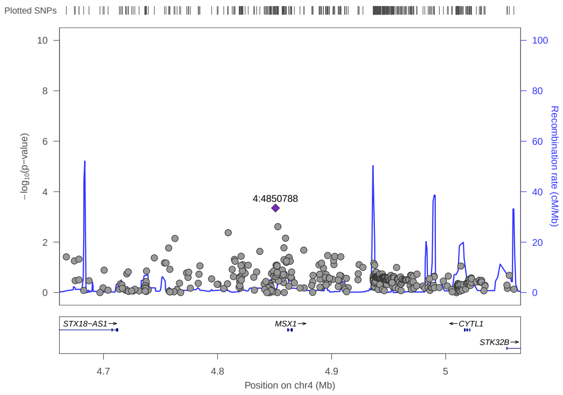


**(d)** *MSX1*

**Figure S18: Regional association plots to investigate if genetic overlap between CL/P & CP is identified by PLACO at some candidate regions based on past literature.** The blue diamond represents the most strongly associated SNP in the region of genetic overlap. For multi-ethnic analyses, there is no unique LD between SNPs and hence no color has been used to represent strength of LD.

#### Supplementary S2

##### Empirical validation of sensitivity and specificity of PLACO.

**Simulation setup:** We simulated two bi-ethnic case-parent trio studies with a total of 2400 complete trios mimicking independent studies of CL/P and CP. We assumed, without loss of generality, that the two ethnic groups have equal sample sizes for a particular OFC subgroup, and considered situations where the OFC subgroups either have comparable/balanced (1:1) or unbalanced (3:1) or heavily unbalanced (7:1) sample sizes. For instance, the unbalanced (3:1) scenario simulates a total of 2400 trios of which 1800 are CL/P trios (900 in each ethnic group), and 600 are CP trios (300 in each ethnic group). Thus, the two studies with comparable (1:1) or unbalanced (3:1) or heavily unbalanced (7:1) sample sizes respectively have sample sizes ($n_{\mathrm{CL/P}}$, $n_{\mathrm{CP}}$) = (1200,1200) or (1800,600) or (2100,300). Our choices of the unbalanced and the heavily unbalanced OFC subgroup sample sizes mimic the GENEVA and the POFC studies respectively (**Table S1**).

We made the two ethnic groups distinct in two ways. First, we assumed that the OFC subgroup prevalence for each ethnic group is different. In particular, we used the cleft prevalence for Western Europe and South Asia[1]. CL/P prevalence for these two ethnic groups are 1.07/1000 and 1.30/1000, while those of CP are 0.59/1000 and 0.30/1000 respectively. Secondly, we assumed the minor allele frequency (MAF) of any given genetic variant is different between ethnic groups (as described next). Recall, the MAFs for the top SNPs from the regions of genetic overlap between CL/P and CP that we identified from the analysis of POFC + GENEVA data are mostly different across ethnic groups.

For the genetic data, we simulated 10 million independent bi-allelic genetic variants in Hardy-Weinberg equilibrium (HWE). We fixed the population-level MAF of one ethnic group at 10%; for the other ethnic group, we randomly selected the MAF between 7% and 13% using a continuous uniform distribution. We assumed the commonly-used additive genetic mode of inheritence, and used the gTDT conditional logistic model[2] as our outcome generative model for a given variant:

$$P\Big(Y_0 = 1 \Big| \{Y_0 + Y_1 + Y_2 + Y_3 = 1\}, \{G_0, G_1, G_2, G_3\}\Big) = \frac{e^{\beta G_0}}{e^{\beta G_0} + e^{\beta G_1} + e^{\beta G_2} + e^{\beta G_3}}$$

where $\beta = \log(\text{RR})$ is the genetic effect of the variant for a given OFC subgroup, $Y_0$ is the disease status of the child ($Y_0 = 1$ in a case-parent trio study), $Y_l$ is the disease status of the $l$-th pseudo-control (takes value 0 for all pseudo-controls, $l = 1, 2, 3$), $G_0$ is the genotype of the child at the variant (coded additively as 0, 1, or 2 here), and $G_l$ is the genotype of the $l$-th pseudo-control. Essentially, $G_1$, $G_2$ and $G_3$ are the possible genotypes at the bi-allelic variant that the child could have inherited from the parents. We emphasize that this generative model for our simulation experiments is distinct from the hierarchical model assumed by PLACO[3]. Since we need multiple independent replicates to assess type I error control and power at stringent error thresholds, we assumed the 10 million genetic variants are independent. Subsequently, we calculated estimated type I error (power) by averaging over the number of independent null (non-null) variants identified as having significant pleiotropic effect on both outcomes at a fixed significance level $\alpha$ (the choice of $\alpha$ is mentioned when presenting results).

Out of the 10 million genetic variants, we assumed 99% variants to be not associated with either of the two OFC subgroups (i.e., $\text{RR}_{\text{CL/P}} = 1$, $\text{RR}_{\text{CP}} = 1$), 0.5% variants to be associated with CP only (i.e., $\text{RR}_{\text{CL/P}} = 1$, $\text{RR}_{\text{CP}} \neq 1$), 0.4% variants to be associated with CL/P only (i.e., $\text{RR}_{\text{CL/P}} \neq 1$, $\text{RR}_{\text{CP}} = 1$), and 0.1% variants to be associated with both (i.e., $\text{RR}_{\text{CL/P}} \neq 1$, $\text{RR}_{\text{CP}} \neq 1$). Thus, our simulated dataset had 9.99 million null variants to estimate type I error and $10,000$ non-null variants to estimate statistical power. Note, we have purposefully simulated a very large number of null variants to enable meaningful type I error comparison between PLACO and other methods at stringent error thresholds typically used in GWAS. The different choices of $\text{RR}_{\text{CL/P}}$ and $\text{RR}_{\text{CP}}$ for simulating our data will dictate the scenarios under which we evaluated type I error and power of PLACO in identifying variants having simultaneous association with CL/P and CP based on multi-ethnic case-parent trios.

**Type I error comparisons:** We considered two primary scenarios: we assumed either (I) fixed genetic effects for both OFC subgroups across variants, or (II) a distribution on the genetic effects of CL/P group and fixed genetic effects for CP. Specifically, for Scenario I we assumed 9.9 million null variants with $\{\text{RR}_{\text{CL/P}} = 1, \text{RR}_{\text{CP}} = 1\}$; $50,000$ null variants with $\{\text{RR}_{\text{CL/P}} = 1, \text{RR}_{\text{CP}} = 1.15\}$; and $40,000$ null variants with $\{\text{RR}_{\text{CL/P}} = 1.15, \text{RR}_{\text{CP}} = 1\}$. For Scenario II, we assumed 9.99 million null variants with $\log(\text{RR}_{\text{CL/P}})$ simulated from a normal distribution with mean 0, standard deviation 0.1, and fixed $\text{RR}_{\text{CP}} = 1$. The choice of this

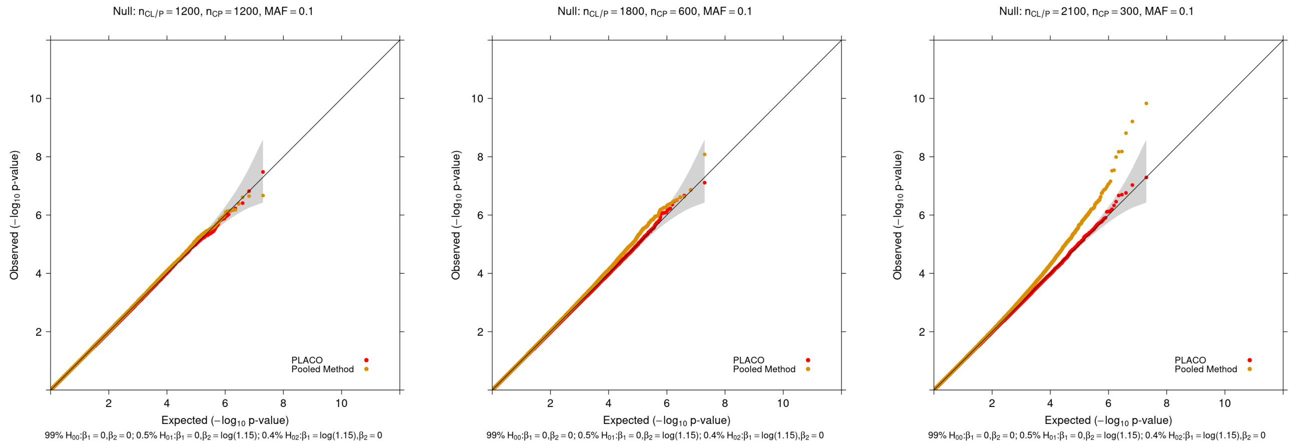


**(a)** MAF 10%; Balanced (1:1)

**(b)** MAF 10%; Unbalanced (3:1)

**(c)** MAF 10%; Heavily unbalanced (7:1)

**Figure S19: Scenario I: QQ plots for null data from two independent bi-ethnic case-parent trio studies of OFC subgroups assuming fixed genetic effects.** Observed($-\log_{10}$p-values) are plotted on the y-axis and Expected($-\log_{10}$p-values) on the x-axis. Type I error performance of tests of simultaneous effect of a genetic variant on both outcomes is based on 9.99 million null variants with genetic effects that are either $\{RR_{CL/P} = 1, RR_{CP} = 1\}$ or $\{RR_{CL/P} = 1, RR_{CP} = 1.15\}$ or $\{RR_{CL/P} = 1.15, RR_{CP} = 1\}$. The gray shaded region represents a conservative 95% confidence interval for the expected distribution of p-values. P-values $\geq 10^{-12}$ are shown here.

distribution for $\log(RR_{CL/P})$ is motivated by the distribution of effect sizes of common variants across many complex human traits[4]. While Scenario I has bulk of the null variants under the global null (i.e., both genetic effects are null), Scenario II ensures that the bulk of null variants is under the sub-null where only one cleft group shows genetic association. Scenario I is simple and straightforward; Scenario II is more realistic.

Further, to evaluate sensitivity (if any) of PLACO's type I error control to varying MAF, we considered a simulation setting with everything the same as before except that the MAF of the variants for one ethnic group is fixed at 20% while that of the other ethnic group is allowed to vary more widely around 20% by simulating its MAF between 5% and 35% using a continuous uniform distribution. To avoid redundancy, we did this only for the more realistic Scenario II.

We compared PLACO with 'pooled method' GWAS analysis[5] previously used to identify risk variants common to both CL/P and CP. The pooled method combined the OFC subgroups together to form a single OFC group. When almost all of the null variants have no effect on either OFC subgroup (Scenario I), both PLACO and the pooled method appear to have controlled type I error for balanced sample sizes of the OFC subgroups (**Figure S19 (a)**). As the OFC subgroups become more and more skewed in terms of sample size (as seen for our

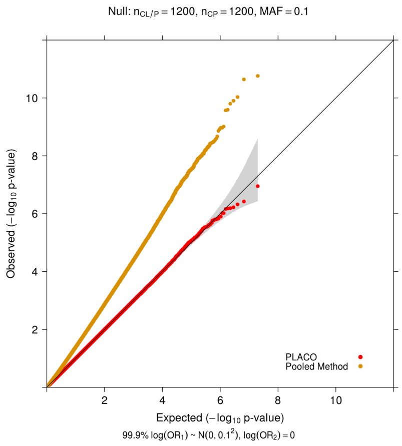


**(a)** MAF 10%;
Balanced (1:1)

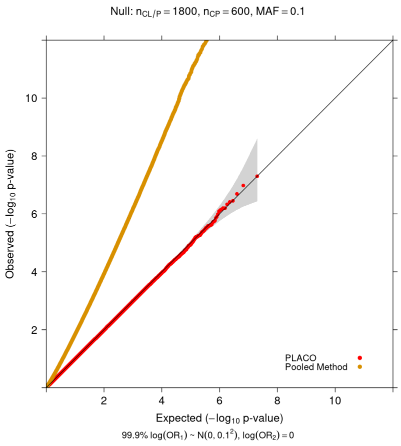


**(b)** MAF 10%;
Unbalanced (3:1)

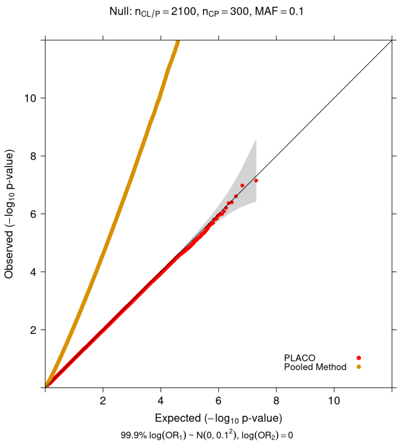


**(c)** MAF 10%;
Heavily unbalanced (7:1)

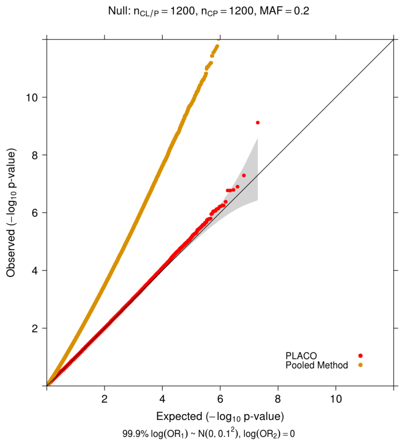


**(d)** MAF 20%;
Balanced (1:1)

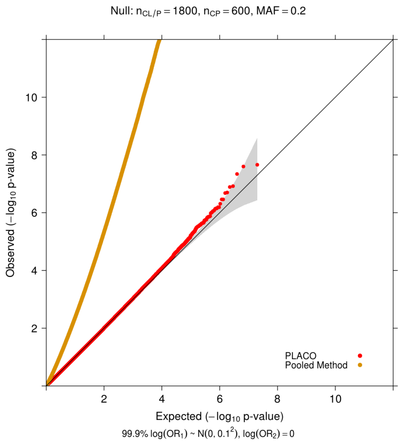


**(e)** MAF 20%;
Unbalanced (3:1)

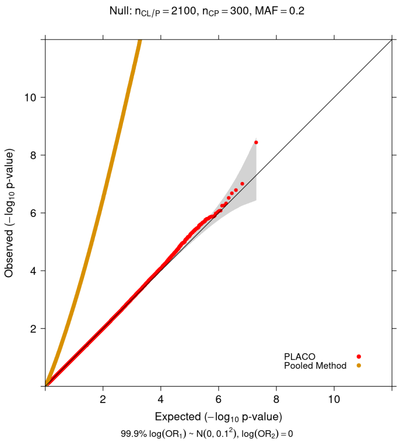


**(f)** MAF 20%;
Heavily unbalanced (7:1)

**Figure S20: Scenario II: QQ plots for null data from two independent bi-ethnic case-parent trio studies of OFC subgroups assuming fixed genetic effects for one trait and random for the other.** Observed($-\log_{10}$p-values) are plotted on the y-axis and Expected($-\log_{10}$p-values) on the x-axis. Type I error performance of tests of simultaneous effect of a genetic variant on both outcomes is based on 9.99 million null variants with genetic effects $\{\log(\mathrm{RR}_{\mathrm{CL/P}}) \sim N(0, 0.1^2), \mathrm{RR}_{\mathrm{CP}} = 1\}$. The gray shaded region represents a conservative 95% confidence interval for the expected distribution of p-values. P-values $\geq 10^{-12}$ are shown here.

GENEVA and POFC studies), the pooled method shows inflated type I error while PLACO maintains appropriate type I error rate even at stringent error levels (**Figure S19 (b)-(c)**). This shows that the 'pooled method' does not necessarily capture only shared signals[6]; it may show spurious signals if sample sizes are widely different for the subgroups (e.g., CL/P group is almost always much larger than CP group).

When a large proportion of null variants have genetic effect on one OFC subgroup only with the genetic effect randomly ranging from being weak to strong (Scenario II), the pooled method shows hugely inflated type I error rate while PLACO still maintains proper type I

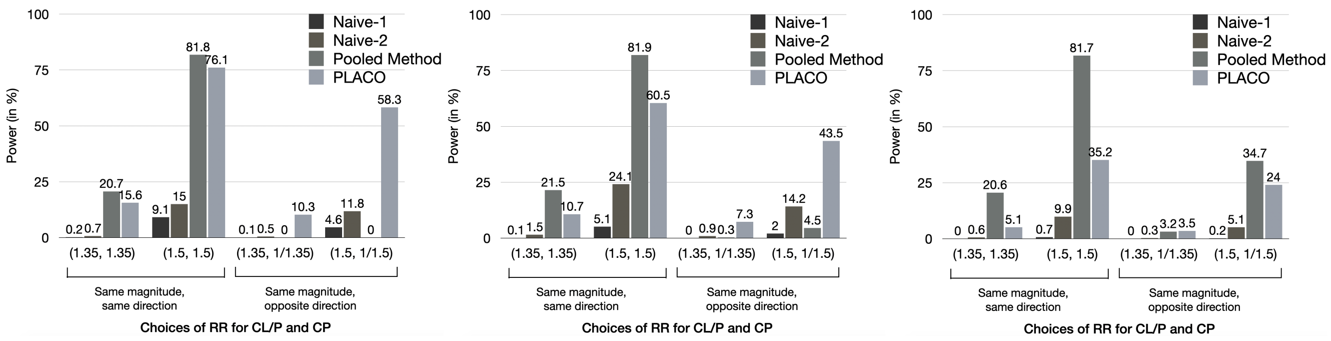


**(a)** MAF 10%; Balanced (1:1)

**(b)** MAF 10%; Unbalanced (3:1)

**(c)** MAF 10%; Heavily unbalanced (7:1)

**Figure S21: Power of PLACO, pooled method, and naive approaches at genome-wide significance level ($5\times10^{-8}$) for varying genetic effects of the two independent bi-ethnic case-parent trio studies of OFC subgroups.** The first naive approach ('Naive-1') declares pleiotropic association when $p_{\mathrm{CL/P}} < 5\times10^{-8}$ and $p_{\mathrm{CP}} < 10^{-5}$, while the second naive approach ('Naive-2') uses a more liberal criterion $p_{\mathrm{CL/P}} < 5\times10^{-8}$ and $p_{\mathrm{CP}} < 10^{-3}$. Note, unlike PLACO, the pooled method lacks type I error control in most scenarios of sample size and/or MAF imbalance, and hence its power should be interpreted with caution.

error control at stringent levels regardless of how skewed the sample sizes are between the two OFC subgroups (**Figure S20 (a)-(c)**). This observation holds true for varying MAFs irrespective of how widely different the MAFs are between the two ethnic groups (**Figure S20 (d)-(f)**). This simulation experiment shows that the 'pooled method' is prone to exhibiting spurious signals if genetic effects exist in one OFC subgroup but not the other. On the other hand, we observe the robustness of PLACO's type I error control to sample size differences between OFC subgroups; moderately strong subgroup-specific effects; and small to large MAF differences between ethnic groups.

**Power comparisons:** For the 10,000 non-null variants in our simulated dataset used to estimate power, we considered different choices of the two relative risks $\mathrm{RR}_{\mathrm{CL/P}}$ and $\mathrm{RR}_{\mathrm{CP}}$ to reflect genetic effects of varying directions and/or magnitudes. For benchmarking, we compared power of PLACO against the pooled method (even though pooled method showed inflated type I error in most of our simulated scenarios). We also included the naive approach of declaring genetic overlap when a variant reaches genome-wide significance for the OFC subgroup with a larger sample size (in our case, CL/P) and reaches a more liberal significance threshold for the other. We used two such naive approaches: one based on criterion $p_{\mathrm{CL/P}} < 5\times10^{-8}$, $p_{\mathrm{CP}} < 10^{-5}$ and the other $p_{\mathrm{CL/P}} < 5\times10^{-8}$, $p_{\mathrm{CP}} < 10^{-3}$ ('Naive-1' and 'Naive-2' respectively in our figures). Regardless of the magnitude and directions of simultaneous association and the sample size differences between OFC subgroups, PLACO shows dramatically improved statistical power to detect common genetic basis compared to the naive approaches (**Figure S21**). Pooled method is slightly more powerful than PLACO in identifying shared risk variants (i.e., variants with genetic effects on the two OFC subgroups in the same direction); however, it especially lacks power to detect variants that increase risk for one OFC subgroup while decreasing risk for the other. Note that for the hugely unbalanced (7:1) sample size scenario, the pooled method appears to be more powerful than PLACO for opposite genetic effects; this behavior is explained by the fact that pooled method is strongly influenced (biased away from the null) by sample size differences between groups, and its type I error control can be particularly bad in this scenario (**Figures S19(c)**, **S20(c)** and **S20(f)**).
